# The effectiveness and cost of integrating pharmacists within general practice to optimize prescribing and health outcomes in primary care patients with polypharmacy: A systematic review

**DOI:** 10.1101/2022.12.15.22283519

**Authors:** Aisling Croke, Karen Cardwell, Barbara Clyne, Frank Moriarty, Laura McCullagh, Susan M. Smith

## Abstract

**Background:** Polypharmacy and associated potentially inappropriate prescribing (PIP) place a considerable burden on patients and represent a challenge for general practitioners (GPs). Integration of pharmacists within general practice (herein ‘pharmacist integration’) may improve medications management and patient outcomes. This systematic review assessed the effectiveness and costs of pharmacist integration.

**Methods:** A systematic search of ten databases from inception to January 2021 was conducted. Studies that evaluated the effectiveness or cost of pharmacist integration were included. Eligible interventions were those that targeted medications optimization compared to usual GP care without pharmacist integration (herein ‘usual care’). Primary outcomes were PIP (as measured by PIP screening tools) and number of prescribed medications. Secondary outcomes included health-related quality of life, health service utilization, clinical outcomes, and costs. Randomised controlled trials (RCTs), non-RCTs, interrupted-time-series, controlled before-after trials and health-economic studies were included.

Screening and risk of bias using Cochrane EPOC criteria were conducted by two reviewers independently. A narrative synthesis and meta-analysis of outcomes where possible, were conducted; the certainty of evidence was assessed using the Grading of Recommendations, Assessment, Development and Evaluation approach.

**Results:** In total, 23 studies (28 full text articles) met the inclusion criteria. In ten of 11 studies, pharmacist integration probably reduced PIP in comparison to usual care (moderate certainty evidence). A meta-analysis of number of medications in seven studies reported a mean difference of -0.80 [-1.17, -0.43], which indicated pharmacist integration probably reduced number of medicines (moderate certainty evidence). It was uncertain whether pharmacist integration improved health-related quality of life because the certainty of evidence was very low. Twelve health-economic studies outlined costs and potential cost-effectiveness.

**Conclusions:** Pharmacist integration probably reduced PIP and number of medications however, there was no clear effect on other patient outcomes; and while interventions in a small number of studies appeared to be cost-effective, further robust, well-designed cluster RCTs with economic evaluations are required to determine cost-effectiveness of pharmacist integration within general practice.

**PROSPERO Registration:** [CRD42019139679](https://www.crd.york.ac.uk/PROSPERO/display_record.php?RecordID=139679)

## Background

Polypharmacy places a considerable burden on both patients and health care providers through an increased risk of PIP, increased treatment burden, adverse outcomes, and medication-related hospitalizations (1). Polypharmacy is typically defined as using five or more regular medications (2). A recent systematic review estimated PIP prevalence in primary care to be 33%, with 7% to 17% of all adverse outcomes related to older persons in primary care (3). Various interventions have been trialled to improve medications optimization, including addressing polypharmacy, PIP and deprescribing (the process of withdrawal, including dose reduction, of an inappropriate medication, supervised by a healthcare professional (4)) with mixed effects being reported (5-7). Interventions with organizational (pharmacist supported interventions), professional and multifaceted approaches may provide modest benefits (5).

While strategies for pharmacist interventions have been found to have a positive effect on medication-related problems in hospital and nursing home settings (8, 9), the evidence base for pharmacist interventions within general practice is varied. Barriers have been identified that reduce the ability of community pharmacists to deliver the most effective care to patients and support GPs; these barriers include lack of integration with the general practice team, time restrictions, poor interprofessional communication, lack of access to patients’ medical histories and health policies which discourage collaborative agreements within primary care settings (10). Therefore, one strategy to address these issues may be pharmacist integration within the general practice team either by co-location (herein ‘co-located integration’) or remotely. The pharmacist may not be present in the same geographical location as the GP but based in a community pharmacy and integrated in terms of a formal pathway for communication of medication review issues with the GP (herein ‘remote integration’).

Co-located integration of pharmacists has been shown to deliver a range of non-dispensing interventions, with medication management reviews being a primary activity (11). Systematic reviews have reported mixed effects for these interventions on medications optimization outcomes such as level of PIP and deprescribing of inappropriate medications (12, 13). However, the PINCER trial in the UK demonstrated that such interventions were effective at reducing medication-related errors (14). Pharmacist integration may also reduce GP workload directly through supports for medication-related administration and management, medications reconciliation following hospital discharge and indirectly though reducing medication related adverse events leading to emergency department attendance and hospitalizations (15). Issues surrounding heterogeneity, study quality and missing data, make conclusions about the effectiveness of interventions difficult to draw (13).

The evidence base to determine whether such interventions are cost effective also requires further study (12). The association between polypharmacy and adverse drug events (ADEs) gives rise to substantial costs to both the healthcare system/health service and patients (16). An estimated 237 million medication errors occur annually in England, with approximately 38% occurring in primary care. Avoidable ADEs resulted in an estimated £96,462,582 cost to the National Health Service (NHS) in 2018 (17). Where interventions in hospital settings involving pharmacist and physician collaboration can result in cost-avoidance (18), there is little evidence regarding cost-effectiveness in primary care settings.

Previous reviews of pharmacist interventions focused solely on co-located integration (12, 13, 15, 19). This paper systematically updated this evidence and reviewed the literature on the effectiveness and cost of pharmacist integration within general practice, to improve prescribing practices and health outcomes for adult patients with polypharmacy in the primary care setting. A secondary aim was to explore and report the domains of integration for these interventions.

## Methods

This systematic review was conducted using Cochrane methodology (20) and reported using the Preferred Reporting Items for Systematic Reviews and Meta-Analyses (PRISMA) guidelines (21). The review was registered on Prospero and a peer-reviewed protocol was published (22).

### Data Sources and Search Strategy

An electronic database search was conducted in 10 databases ([PubMed](https://www.ncbi.nlm.nih.gov/pubmed/), [Cochrane Library](https://www.cochranelibrary.com/), [Cochrane Central Register of Controlled Trials](https://www.cochranelibrary.com/central/about-central), [EMBASE](https://www.embase.com/login), [Web of Science](https://apps.webofknowledge.com/WOS_GeneralSearch_input.do?product=WOS&search_mode=GeneralSearch&SID=D3l81bxsbEG4X11SEIM&preferencesSaved=), [SCOPUS](https://www.scopus.com/search/form.uri?display=basic), [Lilacs](https://lilacs.bvsalud.org/en/) and [CINAHL](https://www.ebscohost.com/nursing/products/cinahl-databases/cinahl-complete)) from inception to end of January 2021 using a combination of free text terms, keywords and Medical Subject Headings (MeSH). No language or date restrictions were applied (see Additional file 1).

The systematic literature search for the health-economic studies was conducted in NHS Economic Evaluations Database (NHS EED) and the Health Technology Assessment (HTA) database, and an economic filter was applied to both PubMed and EMBASE. A combination of free text terms, keywords and MeSH terms were applied as above.

### Eligibility criteria

Studies were included if they met the following inclusion criteria:

#### Participants

Community dwelling patients aged 18 years and over in the primary care setting with polypharmacy. Studies had to have a majority of patients (≥80%) identified as having polypharmacy (using any definition). Only studies conducted in the primary care setting were included. The definition of primary care for this review was; “integrated, easy to access, health care services by clinicians who are accountable for addressing a large majority of personal health care needs, developing a sustained and continuous relationship with patients, and practicing in the context of family and community” (23).

Pharmacists involved in medications optimization roles and co-located or remotely integrated within general practice in the primary care setting. Pharmacist interventions in a nursing home, secondary or tertiary care setting were excluded. Domains of integration were adapted from the framework defined by Walshe and Smith (24), with definitions drawing on a previous systematic review (19), as shown in Table 1. These agreed definitions were that four to six domains of integration indicate robust integration, two to three domains indicate moderate integration, and one domain of integration indicates the minimum level of integration.

**Table 1.** Description of integration domains by Walshe and Smith (19)

| **Dimension** | **Definition** |
| --- | --- |
| Organizational | Pharmacist is physically co-located with the GP or, the intervention is remote but encompassed within the same network |
| Informational | Integration and access to clinical patient systems |
| Clinical | Care delivery to patients and communication with GPs |
| Functional | Capture of other actions taken by pharmacists integrated within GP settings such as medications education or administrative support |
| Normative | Design of intervention in terms of shared goals and visions of activities involved and desired outcomes |
| Financial | Financial implications from internally funded pharmacist interventions |

Text highlighted in bold indicate main column headings

#### Intervention

‘Pharmacist integration’ defined as all types of interventions targeted at patient or prescriber behaviours involving a pharmacist aiming to optimize medications for patients in a primary care setting were considered for inclusion. The relationship between the pharmacist and the GP could be conducted by co-located integration or by remote integration providing the relationship continued for the duration of the intervention.

#### Control

Usual GP care that did not include pharmacist integration.

#### Study design

As per the Cochrane Effective Practice and Organisation of Care (EPOC) study design criteria for effects of interventions (25), we included randomised controlled trials (RCTs), cluster RCTs (cRCTs) non-randomised controlled trials (nRCTs), controlled before-after studies (CBA) and interrupted time series (ITS) studies.

Health-economic studies including comparative resource use studies and health-economic evaluations (cost-effectiveness analysis, cost-utility analysis, cost-minimization analysis, and cost-benefit analysis) were also eligible for inclusion.

#### Outcomes

The primary outcomes for this review included:

- PIP or high risk prescriptions as reported by included studies. Studies reported potentially inappropriate or high risk prescriptions using screening tools such as; Screening Tool of Older Person’s Prescriptions / Screening Tool to Alert doctors to Right Treatment (STOPP/START) and Beers criteria (explicit criteria), or the Medications Appropriateness Index (MAI), Prescribing Appropriateness Index and Drug Burden Index (DBI) (implicit criteria).
- The per-patient number of medications prescribed and change in the number of medications prescribed as reported by included studies. The definition varied across studies (e.g. some may use the number of repeat medications), however where possible we used the number of medications including acute and repeat prescribed medications.

Secondary outcomes included:

- Health-related quality of life (HRQoL)
- Adverse events or harms
- Health service utilization
- Clinical physical outcomes
- Mental health outcomes
- Comparative resource use, costs and cost-effectiveness

### Study Selection and Data Extraction

Citations were downloaded to Endnote (26) and duplicates removed. Titles were screened for clearly ineligible studies by one researcher (AC). Remaining titles and abstracts were independently screened using Rayyan software (27), by at least two of the three members of the review panel (AC, OJ and KC). Full text suitability for inclusion was independently determined by two researchers (AC and KC). Disagreement was managed by consulting a third reviewer (FM).

Data were extracted by two reviewers (AC and KC) on name of first author, year of publication, country of publication, study setting; study population and participant demographics, intervention details and design including framework of integration elements, control, setting details, and outcomes.

### Data Synthesis

Interventions were assessed for the six dimensions of integration dichotomously (yes/no).

Due to the heterogeneity relating to the wide variation in participants, interventions and outcomes assessed, the main synthesis of the results is presented narratively. A meta-analysis using inverse variance with random effects statistical models for continuous variables with mean difference effect measures was conducted for one of the primary outcomes, number of medications, using data from eligible RCTs only. Heterogeneity was assessed using the I^2^ statistic, the percentage of variability in the estimates due to heterogeneity, and interpreted as per the Cochrane Handbook, 0% to 40%: might not be important; 30% to 60%: may represent moderate heterogeneity; 50% to 90%: may represent substantial heterogeneity; 75% to 100%: considerable heterogeneity (20).

Subgroup analysis was based on location of intervention (co-located vs remote integration) and degree of polypharmacy. It was not possible to conduct subgroup analysis based on age of patients given the data presented in studies.

The costs for health-economic studies were inflated to 2021 prices using the Consumer Price Index (CPI) for each individual country and converted to euro (where appropriate) using the purchasing power parity (PPP) indices by the Organization for Economic Co-operation and Development (OECD).

Sensitivity analyses for estimates of effect size and determinants were not assessed owing to limitations in the data reported for studies.

### Risk of Bias

The risk of bias in all included effectiveness studies was assessed by two reviewers (AC and KC) using standard EPOC criteria (25) including the following domains: allocation (sequence generation and concealment); baseline characteristics; incomplete outcome data; contamination; blinding; selective outcome reporting; and other potential sources of bias. Robvis online software was used to generate risk of bias figures (28). The health-economic studies were assessed for methodological quality using the Consensus on Health Economic Criteria (CHEC) (29) list by one reviewer (AC).

### Assessing quality of included studies

The certainty of evidence for five critical and important outcomes was assessed using the Grading of Recommendations Assessment, Development and Evaluation (GRADE) criteria and [GRADEPro software](https://community.cochrane.org/help/tools-and-software/gradepro-gdt) and judgements are presented in a ‘Summary of findings’ (SOF) table (30). The five outcomes assessed were PIP, number of medications, ADEs, HRQoL and mortality. These outcomes were selected in accordance with the Core Outcome Set (COS) for Trials Aimed at Improving the Appropriateness of Polypharmacy in Older People in Primary Care (31).

## Results

### Search results

A total of 26,887 articles were retrieved up to the end of January 2021. Full texts of 207 articles were assessed for eligibility and 28 full texts were included in the systematic review (Figure 1).

**Figure 1.** Prisma flow chart for included studies

### Characteristics of included studies

#### Included studies, participants and outcomes reported

A total of 23 studies were reported across 28 articles. Seven studies were conducted in North America (32-38), three in the United Kingdom (UK) (39-41), ten in other European countries (42-51), and three in New Zealand or Australia (52-54) (Table 2). The age range of the 23,516 included participants was 1 to 102 years of age (one study reported ages from 1 to 102, however median age in that study was 65 years so study was included (39)) and the number of medications prescribed per person ranged from 3 to 27. In addition to the broad review inclusion criteria of polypharmacy, three studies had further inclusion criteria relating to high frequency of daily dosing (≥12 doses per day) and drugs that required monitoring (33, 37, 53). Two studies included participants with more than three current disease states (33, 37) and one study included patients with 50 or more prescriptions filled in the previous year (100% correlation to 5 or more medications) (35).

Formal training qualifications or requirements for the pharmacists were outlined in five studies (32, 41, 46, 51, 52), one study detailed training provided to GPs and pharmacists by the study team (54) and two studies stated prior experience of clinical training of pharmacist(s) was required (40, 42).

Three of the 23 studies had cluster randomised designs (48, 49, 54), 17 studies had an individual patient randomised design, one study had a non-randomised design, and one adopted a controlled before-after design. Eleven studies recorded PIP outcomes (34, 37, 39, 40, 42-45, 47, 51, 52), and nine studies reported on differences in number of medications between pharmacist integration and usual care groups (32, 34, 37, 41, 42, 46-48, 55). Review secondary outcomes included 15 studies (16 articles) which reported on HRQoL; nine studies reported Short Form 36 Health Survey Questionnaire (SF-36) (33, 34, 36, 37, 40, 48, 52-54) and six studies (seven articles) reported EuroQol-5D (EQ5D) (41, 42, 45, 46, 49, 50, 55). Five studies reported on ADEs (35, 37, 45, 50, 54), 10 studies reported on health service utilization (37, 38, 40-42, 45, 48, 50, 51, 53) and three studies reported on clinical physical outcomes (33, 37, 43). No study reported mental health outcomes. Government bodies, university departments, or professional bodies funded all studies.

#### Interventions and comparators

All studies reported a pharmacist conducting a medication review with patients to optimize prescribing, nine reported co-located integration (32-35, 37, 39, 40, 51, 53) and 14 studies reported remote integration (36, 38, 41-50, 52, 54). Intervention duration ranged from 60 days to 24 months.

Eighteen studies compared pharmacist integration with a comparator described as ‘usual care’ (32-40, 42, 43, 47-51, 53, 54). In all, ‘usual care’ was considered to be standard best practice with no pharmacist integration. Of the 23 included studies, five studies adopted a wait-list control (41, 44-46, 52)

*<Table 2. Characteristics of included effectiveness studies>*

### Characteristics of included health-economic studies

Twelve health-economic studies were included in the review, four were conducted in the US (32, 35, 56, 57), three were conducted in Spain (55, 58, 59), one was conducted in multiple EU countries (48), one in the UK (40), one in Canada (36), one in the Netherlands (51) and one in Australia (54). All health-economic studies were further analyses of 11 primary studies already included in the systematic review (Table 3). Three studies presented cost-effectiveness analyses; two were cost-effectiveness analyses (CEA) (54, 56) and one was a cost-utility analysis (CUA) (55). One study presented a cost-benefit analysis (58) and nine studies detailed cost (32, 35, 36, 40, 48, 51, 57-59).

The age of participants ranged from 25 to 100 years. Six studies outlined co-located integration (35, 40, 51, 56-58) and five studies (six articles) investigated remote integration (32, 36, 48, 54, 55, 59). All studies adopted a third-party payer perspective. Four studies adopted a 12-month time horizon (55, 56, 58, 59) and one study detailed costs for 12 months before and after the intervention (57). Seven studies did not state a specific time horizon but outlined that data was collected in line with intervention duration (32, 35, 36, 40, 48, 51, 54).

*<Table 3. Characteristics of included economic evaluations>*

### Domains of integration in included effectiveness studies

Five studies had robust integration (organizational, informational, clinical, and financial) (33, 34, 36, 37, 43). Seventeen studies had moderate integration (32, 35, 38-42, 44-47, 49-54). One study had a minimum level of integration (48). Details of domains of integration associated with different studies are outlined in Table 2 and summarized in Additional file 2.

### Risk-of-Bias Summary

For included RCTs, there was low or unclear risk of bias (Figure 2) across the majority of domains. Most studies however had a high risk of bias in relation to protection against contamination. The most common issue leading to a judgement of unclear risk of bias was lack of clarity around blinding of participants. There was high risk of bias for all nRCTs due to limitations in randomization and allocation concealment and a high risk of bias due to knowledge of allocation across all studies (Figure 3). The full risk of bias assessment for all outcomes is available in Additional file 3.

**Figure 2.** EPOC Risk of Bias assessment for RCTs

**Figure 3.** EPOC Risk of Bias assessment for nRCTs

The full text articles related to health-economic studies were assessed for risk of bias. The quality was varied across the included health-economic studies (Figure 4). Missing data was an issue across all studies which did not allow for an estimation of risk of bias in this review. Uncertainty about the rigour of outcomes reporting and sensitivity analyses were also noted.

**Figure 4.** CHEC List for assessing methodological quality of health-economic studies

### Certainty of the evidence

The outcomes included in the SoF Table include the review primary outcomes, and important outcomes identified in the COS which were aligned with the outcomes of this review (See Table 4). In general, the majority of included studies were RCTs and as such, GRADE assessment for certainty of evidence was limited to that study design (30). Of the 23 included studies, 21 were based on an RCT design.

The certainty of evidence relating to the impact of pharmacist integration on PIP was moderate. Studies were downgraded for serious concerns relating to risk of bias. The certainty of evidence for number of medications was moderate due to serious concerns relating to risk of bias and limited to the seven studies included in the meta-analysis. The certainty of evidence for HRQoL was very low due to serious concerns relating to risk of bias, inconsistency of results and imprecision of results. Certainty of evidence for ADEs and mortality was low due to serious concerns relating to risk of bias and imprecision of results and very serious concerns relating to imprecision of results respectively. Economic outcomes were not included in the SoF table.

*<Table 4. Summary of findings table>*

### Primary Outcomes

#### Potentially inappropriate prescribing

Eleven of the 23 studies evaluated effects of pharmacist integration on a range of PIP indicators. Ten were RCTs with moderate certainty of evidence (34, 37, 40, 42-45, 47, 52). Heterogeneity in terms of reported outcomes dictated a narrative synthesis of results. Six of the 11 studies utilised validated screening tools. Three studies used the MAI (34, 37, 52), two of which (34, 52) reported significant changes favouring pharmacist integration (Additional file 4). Two studies reported the Drug Burden Index (DBI) (45, 51) and reported a reduction in the DBI favouring pharmacist integration, significance not reported. One study used the STOPP/START criteria and reported significant improvements in PIP favouring pharmacist integration (42).

Of the remaining five studies, two studies used a structural assessment by Cipolle et al (60) which is an assessment according to a rational order of indication, effectiveness, safety and compliance (43, 44), and three studies used locally defined drug related problems (DRPs) (39, 40, 47) (Additional file 4). Overall, four of these five studies reported an improvement in PIP for pharmacist integration (39, 40, 44, 47) with one study reporting significantly less pharmaceutical care issues (PCIs) for pharmacist integration groups in comparison to usual care (21.2% and 60.7% respectively, RR, 0.35 (95% CI 0.31 – 0.39)) (40), three studies did not report significance (39, 44, 47) and one study favoured usual care (43).

#### Number of medications

Nine studies reported on per-patient differences in number of medications at study endpoint (Additional file 5) (32, 34, 37, 41, 42, 46-48, 55). Direction of effect favoured pharmacist integration in all but one of the studies. Seven of these studies could be included in a meta-analysis (Figure 5) which indicated pharmacist integration probably reduced mean number of medications in comparison to usual care (mean difference -0.80 [95% CI - 1.17 to -0.43]). There was moderate heterogeneity in the reported results as indicated by the I^2^ statistic of 57%.

**Figure 5.** Meta-analysis of number of medications

### Secondary Outcomes

#### HRQoL

Fifteen studies (16 articles) reported on HRQoL, two studies (three articles) reported some improvement (49, 53, 55), three reported mixed effects (33, 46, 52) and 10 studies reported no difference between groups (Additional file 6) (34, 36, 37, 40-42, 45, 48, 50, 54). Of the two that reported an improvement, one study (two articles) reported a mean difference in utility score (SD) of 0.0550 (0.01) (95% CI 0.0306 to 0.0794) in favour of pharmacist integration (49, 55); and the other reported significant improvements in mental health and vitality favouring pharmacist integration but did not provide data on other domains (53). Of the three with mixed effects, one study favoured usual care with reported improvements in HRQoL in two domains; emotional role and social functioning but provided no further data (52). One study only reported an improvement in the VAS score (46) and the other study did not report usual care group data thus an estimate could not be made (33).

Six of nine studies using the SF-36 (34, 36, 37, 40, 48, 54) showed no significant difference between pharmacist integration and usual care across all eight domains. Two studies reported some mixed effects across the different SF36 domains (33, 52). The remaining study reported an improvement (53). Four of six studies used the EQ5D and reported no significant difference between groups at follow-up (41, 42, 45, 50). The other two studies (three articles) reported significant differences though in Verdoorn et al this was only in the EQ5D VAS and not in the index score (46, 49, 55).

#### ADEs

Five studies reported on ADEs (Additional file 7). (35, 37, 45, 50, 54) Overall three studies reported a decrease in ADEs in the pharmacist integration group versus usual care, though these were not significant differences. Of the remaining two studies, one reported improved adverse effect scores and symptoms in the pharmacist integration group (p=0.024) (35), and the remaining study reported no significant difference between groups (50).

#### Health service utilization & Mortality

Ten studies reported on health service utilization (Additional file 8). (37, 38, 40-42, 45, 48, 50, 51, 53). Seven studies reported no significant difference between groups at follow-up (38, 41, 42, 45, 48, 50, 53). Of the three studies which reported a reduction in hospitalizations associated with pharmacist integration, one reported a reduction of hospitalizations of 47% in emergency admissions however the reported numbers were deemed too small for meaningful statistical analysis (40). The other two studies reported a reduction in hospitalizations; one study reported the adjusted rate ratio for medication-related hospitalizations in the pharmacist integration group compared to usual care as 0.68 (95% CI 0.57–0.82) (51). The remaining study reported a significant difference in reported hospitalizations over the course of the intervention (p=0.003) which favoured pharmacist integration.

Two studies reported on mortality and no effect was found in either study. (42, 45)

#### Clinical physical outcomes

Three studies reported on clinical physical outcomes (33, 37, 43). No significant changes were noted in body mass index (BMI) or renal function (43). In one study, all patients in the pharmacist integration group had international normalised ratios (INRs) within the targeted range, compared with 25% of usual care patients (37) (Additional file 9). Two studies reported significant improvements in blood pressure (BP) management which favoured pharmacist integration (37, 43). Three studies reported mixed results for glycaemic control, two reported no effect on glycosylated haemoglobin (HbA1c) levels (33, 43) and one reported significantly more patients in the pharmacist integration group had achieved their therapeutic goal (37). These three studies all reported significant improvement in lipid profiles in the pharmacist integration group versus the usual care group (33, 37, 43).

### Results of economic studies

Twelve health-economic studies were included. Two studies were CEAs (56); one was a CUA (55). The CUA (cost year 2014, Spanish jurisdiction) reported that the probability of the medication review with follow-up (MRF) service being cost effective, compared with usual care, was 100% when the willingness to pay threshold ranged from €30,000 to €45,000 per quality adjusted life year (QALY) (Additional file 10) (55). One CEA (cost year 1991, US jurisdiction) reported a cost-effectiveness incremental ratio of €9.55 per one unit change in MAI (56). The second CEA was based on analysis on cost savings. The incremental cost per ADE avoided was €74.18, the incremental cost per case of improvement in severity of illness (as measured by the Duke’s Severity of Illness Visual Analogue Scale) was €69.88 (54).

Nine studies considered costs (32, 35, 36, 40, 48, 51, 57-59). Two articles reported on the same study (55, 59), the CUA (55) as outlined above and the other study (cost year 2014, Spanish jurisdiction) detailed the costs and potential price of a (MRF) service and found that mean initial investment per pharmacy was €5899.92 and mean annual maintenance costs of €3940.13 (59). The potential service price ranged from €304.37 to €806.52 per patient per year (59). One study (cost year 2012, Spanish jurisdiction) reported a non-significant reduction in drug expenditure in the pharmacist integration group at follow-up of €321.43 (95% CI 233.77– 409.79) in comparison to usual care €232.94 (95% CI 141.64 – 323.15) (p=0.171) (58). The estimated return on investment of pharmacist integration (control €0 per scenario) ranged from €2.34 to €3.27 per patient per year based on sensitivity analysis (basal, optimistic, and conservative scenarios) (58). A cost analysis study (cost year 1991, US jurisdiction) reported a non-significant difference in change of mean total costs (cost of clinic visits, medications, hospitalizations, and laboratory tests) of -€446.36, p=0.06 (57). Five further cost analysis studies reported no difference in costs between pharmacist integration and usual care groups at follow-up (35, 36, 40, 48, 51). Two studies reported no significant difference between pharmacist integration and usual care in relation to healthcare utilization costs (36, 51). One study reported that total cost savings in the pharmacist integration group of €287.93 was significantly higher than the increase of €1295.93 cost observed in the usual care group (pharmacist integration total cost avoidance €1588.39) (32).

### Sub-group Analysis of Primary Outcomes

Subgroup analysis was based on location of intervention (remote vs co-located integration) and degree of polypharmacy. For PIP, six of the studies investigated remote integration and five investigated co-located integration. All of the studies that examined remote integration favoured pharmacist integration (42-45, 47, 52); one reported a non-significant improvement in the pharmacist integration group (45), the other five studies reported significant changes favouring pharmacist integration. Of the co-located integration studies, 80% favoured the pharmacist integration group (34, 37, 39, 40) and 20% reported mixed results (51). Overall, studies that investigated remote integration favoured pharmacist integration more than those that investigated co-located integration.

Subgroup analysis based on the degree of polypharmacy found that for per-patient number of medications, six studies investigated remote integration (41, 42, 46-48, 55) and three studies investigated co-located integration (32, 34, 37). 83.3% of studies that investigated remote integration favoured pharmacist integration for reducing number of medications (41, 42, 46, 47, 55) and 16.7% found no difference between pharmacist integration and usual care groups (48). All studies that investigated co-located integration favoured pharmacist integration group for reducing the number of medications. Both co-located and remote integration demonstrated a mix of significant and non-significant results which makes estimation of effect difficult.

## Discussion

This review identified 23 studies of the effectiveness of pharmacist integration and eleven of the effectiveness studies reported a health-economic study, with four reported separately in five publications. Across the included studies, there was heterogeneity in terms of medications optimization interventions and health outcomes reported. Overall, ten of 11 studies reporting on PIP outcomes reported pharmacist integration probably reduced PIP (moderate certainty evidence), however one study favoured usual care. A meta-analysis of seven RCTs showed that pharmacist integration can probably reduce the number of medications a patient is prescribed in comparison to usual care (moderate certainty evidence).

Overall, our findings indicate that pharmacist integration may improve patient medications management, but it is uncertain if pharmacist integration improves HRQoL as the certainty of evidence is very low. The majority of included studies reported a decrease in PIP or drug-related problems, a small reduction in the number of medications and a potential reduction in ADEs. All interventions in the 23 included studies involved medication review, only one study included an additional intervention aimed at quality improvement in the practice (51). Our stated inclusion criteria did not stipulate this, although it does logically follow that pharmacist interventions would follow a medication review model given their area of expertise. Patients with polypharmacy are at an increased risk of PIP and ADEs and medication review offers a structure by which these elements can be identified and addressed where appropriate. All studies demonstrated a degree of integration, the majority demonstrated moderate integration across three domains, the most common being organizational, informational, and clinical (32, 35, 38-42, 44-47, 49-54).

Our review findings are in keeping with previous broader systematic reviews, which suggest that pharmacist integration within general practice probably reduce the number of medication-related problems and improve appropriateness of prescribing (5, 13, 14, 61). Previous studies have reported conflicting data in terms of the effect of pharmacist integration on the number of medications (13). This current review found that pharmacist integration probably resulted in a reduction in the number of medications prescribed; this correlates with some published studies (62-64) and conflicts with others (13) which looked at co-located integration only.

Included studies used a variety of screening tools, some were validated tools such as the MAI and STOPP/START criteria and some were locally defined DRPs and pharmaceutical care issues (PCIs). It is likely that many of the criteria used in validated screening tools overlapped with the locally defined protocols. PIP is identified using a variety of implicit and explicit screening tools with the aim of reducing medication harms. A retrospective cohort study identified a 20% reduction in potentially inappropriate medications (PIMs) following pharmacist intervention (64) and evidence from other reviews would suggest that pharmacist interventions targeting PIP may be associated with an improvement in prescribing appropriateness (65) which agrees with the positive trend found in this review.

We based our main outcomes as reported in the GRADE SoF tables, on the COS (31) designed by Rankin et al. The COS is a valuable tool in providing a structure for PIP to be measured in a more consistent manner across studies and enables more robust analysis of available evidence. Given the heterogeneity of outcomes and outcomes measures, reported in this review, more robust evidence is required, and though our review suggests a positive impact, this is based on moderate certainty evidence. Although COSs refer to the importance of assessment of appropriateness, using a screening tool like STOPP or MAI, there was heterogeneity found in terms of the screening tools used and reporting of results in this review which resulted in less robust evidence.

Consistent with current evidence (66, 67) it was uncertain whether pharmacist integration improved HRQoL as measured by the SF-36 and EQ-5D (very low certainty evidence). These measures might not be the most sensitive to the changes in HRQoL that improved medication management may produce. The length of follow-up of included studies may not have been sufficient for the effects of pharmacist integration on HRQoL to manifest. Nonetheless, HRQoL is an important outcome to consider, highlighted by its inclusion in the COS for interventions relating to polypharmacy (31) and it is also necessary to support economic evaluations. The quality of evidence in the trials reporting on HRQoL was very low with critical data needed for determining risk of bias often missing as has been previously reported (13, 15, 65).

Our review suggested that co-located or remote integration likely caused no harm. Interventions were shown to either decrease hospitalizations and emergency admissions or reported no differences between pharmacist integration and usual care and no effect was shown on mortality. However, most studies would have been underpowered to detect rarer adverse outcomes, one study reported ADEs as a primary outcome (37) but no power calculation was reported which likely meant that outcomes reported were underpowered. The findings of our review regarding ADEs and harm are consistent with evidence provided by previous studies (12). One RCT examined the effect of a monitoring plan for medication-related ADEs, which resulted in a decrease in delirium, hospitalization and mortality in the care home setting (68). Other studies have reported multifaceted approaches may reduce PIP (5) whilst others reported uncertainty surrounding reducing PIP (6). There is a paucity of evidence in relation to the effect of pharmacist integration on ADEs in the primary care setting as most are conducted in the secondary or care home settings. While we found no evidence of harms as a result of pharmacist integration, this must be interpreted with a degree of caution.

Previous reviews have suggested that pharmacists in general practice can have positive impacts on clinical outcomes with one systematic review reporting a significant reduction in HbA1c between pharmacist integration and usual care (mean difference −0.88%, 95% CI −1.15% to −0.62%, p< 0.001) (12). The current review found mixed effects on HbA1c levels. Results for interventions on dyslipidaemias indicate significant improvements in lipid profiles which is in agreement with previous systematic reviews (69). Similarly, for BP measurements, pharmacists can improve achievement of target levels, however a previous review found mixed results (70).

The majority of studies involved remote integration (36, 38, 41-50, 52, 54), although results were consistent with those that were co-located. Previous studies have found that the extent of pharmacist integration within a general practice positively influences patient care, this review found that full integration is not required for positive patient outcomes, however this is likely influenced by the fact that even in the remote interventions there were clear structures for pharmacist and GP follow-up and face-to-face communications (19).

There was substantial heterogeneity in the types, results, and quality of included economic evaluations as shown in the risk of bias assessment as set out by the CHEC criteria. While nine studies considered costs and outcomes (32, 35, 36, 40, 48, 51, 57-59), two studies reported intervention cost-effectiveness (54, 56) and a third considered a CUA (55). There was insufficient evidence in this review to determine whether pharmacist integration within general practice is cost-effective. Other studies have reported that interventions delivered at community pharmacies for adults with or at risk of developing acute illness and medical emergencies appear to be cost effective (71). Six studies reported non-significant cost differences with pharmacist integration (35, 36, 40, 48, 51, 57), however costing studies are a useful tool when planning for service provision with one study reporting significant cost savings (32). The CUA reported 100% willingness to pay between €30000/QALY and €45000/QALY, the upper limit of which is comparable to a threshold used by the health payer in Ireland within the drug-reimbursement decision making processes. However, transferability of this CUA to Ireland has not been investigated. Cost-effectiveness evaluations are generally not transferable across jurisdictions given differing methodological requirements and decision-making criteria (72).

### Strengths and limitations

This systematic review involved a comprehensive search of databases with the search design being aided by expert librarians. We included remote and co-located interventions to get a comprehensive overview of interventions where the pharmacist and the GP work together to improve medications management and patient outcomes. No language limits were applied to ensure that all relevant studies were captured during the search process. While the analyses were predominantly narrative, there were sufficient data in the included studies to allow for a meta-analysis of the impact of pharmacist integration on the number of per-patient medications prescribed.

This review included RCTs and other quasi-experimental designs in line with EPOC criteria, which ensured we only included robust study designs as smaller uncontrolled studies produce unreliable estimates of effectiveness. No contact was made with included study authors to resolve unclear information when judging risk of bias which may have led to some studies being downgraded. Baseline imbalances and a lack of allocation concealment could have had significant impacts on reported outcomes.

### Implications for practice and future research

International policy supports pharmacists’ integration within general practice. This review provides further evidence to inform policy in this area. Co-located or remotely integrated pharmacists probably improve PIP and reduce the number of per-patient medications.

In line with the findings of other studies, we concluded that further high-quality economic evaluations should be conducted alongside interventional trials. There is some existing evidence to suggest that pharmacist interventions are cost effective in the primary care setting (71) but we did not find sufficient economic analyses to support this. Evidence suggests that pharmacist integration may positively impact clinical outcomes but this is based on a small number of studies.

A cluster RCT study design should be considered to reduce the risk of bias in future studies where interventions are targeting health professionals providing care for both pharmacist integration and usual care patients (73). Future studies involving pharmacist integration should be powered to assess patient-reported and clinical outcomes, particularly for adverse events and harms. To reduce heterogeneity future studies should report on standardised outcomes, using the COS developed by Rankin et al (31). Currently, as there are no standardised approaches to outcome measurement, synthesising the evidence is challenging owing to the heterogeneity of reporting.

## Conclusions

This review found that pharmacist integration probably reduces PIP and number of medications (moderate certainty evidence). Pharmacist integration may reduce ADEs (low certainty evidence) and make little or no difference to mortality (low certainty evidence) and reported uncertainty whether HRQoL improves (very low certainty evidence). Larger and longer term studies may be needed to explore the impact of pharmacist integration on patient health outcomes, healthcare utilization and costs.

## Supporting information

PRISMA Checklist

## Data Availability

The datasets supporting the conclusions of this article are available in the Open Science Framework repository, The effectiveness and cost-effectiveness of integrating pharmacists within general practice to optimise prescribing and health outcomes in primary care patients with polypharmacy: A systematic review. Extended Data.

https://doi.org/10.17605/OSF.IO/RSWJT

## List of Abbreviations

(ARR): Absolute risk reduction
(ADEs): Adverse drug events
(ATC): Anatomical Therapeutic Chemical
(BP): Blood pressure
(BMI): Body mass index
(EPOC): Cochrane Effective Practice and Organisation of Care
(CI): Confidence intervals
(CHEC): Consensus on Health Economic Criteria
(CPI): Consumer Price Index
(CBA): Controlled before-after trials
(COS): Core Outcome Set
(CEA): Cost-effectiveness analysis
(CUA): Cost-utility analysis
(DBI): Drug Burden Index
(DRPs): Drug related problems
(DUSOI-A): Duke’s Severity of Illness Visual Analogue Scale
(EQ5D): EuroQol-5D
(GPs): General practitioners
(GRADE): Grading of Recommendations Assessment, Development and Evaluation
(HBa1C): Haemoglobin A1c
(HR): Hazard ratio
(HTA): Health Technology Assessment
(HRQoL): Health-related quality of life
(INRs): International normalised ratios
(ITS): Interrupted-time-series
(LDL): Low-density lipoprotein
(MeSH): Medical Subject Headings
(MRF): Medication review with follow-up
(MAI): Medications Appropriateness Index
(MRP): Medication-related problem
(MTM): Medications therapeutic management
(MDT): Multi-disciplinary team
(NHS): National Health Service
(NHS EED): NHS Economic Evaluations Database
(nRCTs): Non-randomised controlled trials
(NNT): Number needed to treat
(OECD): Organization for Economic Co-operation and Development
(PROMs): Patient reported outcome measures
(PCIs): Pharmaceutical care issues
(PDTP): Potential drug therapy problem
(PIMs): Potentially inappropriate medications
(PIP): Potentially inappropriate prescribing
(PRISMA): Preferred Reporting Items for Systematic Reviews and Meta-Analyses
(PCT): Primary care trust
(PPP): Purchasing power parity
(QALY): Quality adjusted life year
(RCTs): Randomised controlled trials
(STOPP/START): Screening Tool of Older Person’s Prescriptions / Screening Tool to Alert doctors to Right Treatment
(SF-36): Short Form 36 Health Survey Questionnaire
(SOF): Summary of findings’
(UK): United Kingdom
(VAS): Visual analogue scale

## Declarations

### Ethics approval and consent to participate

Not applicable

### Consent for publication

Not applicable

### Availability of data and materials

The datasets supporting the conclusions of this article are included in its additional files and are available in the Open Science Framework repository, The effectiveness and cost-effectiveness of integrating pharmacists within general practice to optimise prescribing and health outcomes in primary care patients with polypharmacy: A systematic review. Extended Data. [DOI 10.17605/OSF.IO/RSWJT, https://doi.org/10.17605/OSF.IO/RSWJT]

This project contains the following extended data:

- PubMed Search Strategy for Effectiveness of integrating pharmacists within general practice systematic review.docx (PubMed search strategy)
- Data extraction pilot template
- Risk of Bias tables
- GRADE Assessment Sheets
- Data extraction sheets
- PRISMA checklist for “The effectiveness and cost-effectiveness of integrating pharmacists within general practice to optimise prescribing and health outcomes in primary care patients with polypharmacy: A systematic review.”

Data are available under the terms of the [Creative Commons Attribution 4.0 International Public License](https://creativecommons.org/licenses/by/4.0/) (CC-By Attribution 4.0 International).

### Author’s contributions

AC conceived of the idea for the systematic review, designed the review, conducted the review, drafted the work, approved the submitted version and agreed to be personally accountable for their own contributions.

KC conducted acquisition and analysis of data, approved the submitted version and agreed to be personally accountable for their own contributions.

BC was involved in review design, interpretation of data, revised the work, approved the submitted version and agreed to be personally accountable for their own contributions.

FM was involved in review design, interpretation of data, revised the work, approved the submitted version and agreed to be personally accountable for their own contributions.

LMcC interpretated data, revised the work, approved the submitted version and agreed to be personally accountable for their own contributions.

SMS was involved in review design, interpretation of data, revised the work, approved the submitted version and agreed to be personally accountable for their own contributions.

## Additional files

Additional file 1.docx, PubMed search strategy; sample search

Additional file 2.docx, Domains of integration; summary table

Additional file 3.docx, RoB Assessment for individual outcomes and health-economic studies; 1. PIP, 2. Number of medications, 3. HRQoL, 4. ADRs, 5. Mortality, 6. CHEC list for health-economic studies

Additional file 4.docx, Medication related problems; summary of studies which evaluated PIP

Additional file 5.docx, Difference in number of medications; summary of studies that evaluated number of medications

Additional file 6.docx, HRQoL; summary of studies that evaluated HRQoL using SF-36 and EQ5D

Additional file 7.docx, Change in reported ADEs; summary of studies that evaluated ADEs

Additional file 8.docx, Health service utilization; summary of studies that evaluated health service utilisation

Additional file 9.docx, Clinical physical outcomes; summary of studies that evaluated clinical physical outcomes

Additional file 10.docx, Health-economic studies; summary of results of included health-economic studies

## Acknowledgements

Paul J Murphy MLIS, Information Specialist, Royal College of Surgeons Ireland Library, 26 York Street, D02 YN77. Advised on search strategies.

Grainne McCabe, Scholarly Communications & Research Support Officer, Royal College of Surgeons in Ireland Library, 26 York Street, D02 YN77. Advised on search strategies.

Oscar James, Research Assistant. Completed title and abstract review.

