## Supplementary material for "The effectiveness and cost of integrating pharmacists within general practice to optimize prescribing and health outcomes in primary care patients with polypharmacy: A systematic review": Table 2

### Table 2 Characteristics of included effectiveness studies

| **Author (year,**  **Country)** | **Participants**  **(Patients and healthcare professionals (HCPs))** | **Intervention type, Control group,**  **Domains of integration, Duration of intervention** | **Outcome measures** | **Results primary outcome** |
| --- | --- | --- | --- | --- |
| **Randomised Controlled Trials** | | | | |
| Britton (32)  (1991, USA) | *Total, n:* 572  *Age:* NR  *Number of medications:* *Intervention:* 8.72 $\pm$3.54  *Control:* 8.52 $\pm$3.47  *HCPs:* NR | *Intervention type:* Pharmacist reviewed patient treatment and cost. Medication profile review form attached to each patient file for review by GP. Post GP consult, the pharmacist reviewed files for no. meds, meds changes and compliance.  *Control group:* usual GP care.  *Domains of integration:* Organizational, informational, and clinical domains (moderate).  *Duration of intervention:* 3 months. | Primary  Change in number of medications  Change in cost due to no. of medications  Secondary  Discontinued medications no. and cost  Medications added  Dose change  Cost | *Number of medications at follow-up (mean standard deviation (SD)):*  *Intervention:* -0.21 (1.43)  *Control:* +0.48 (1.18)  P<0.001 |
| Bryant (52) (2011, New Zealand) | *Total, n:* 493  *Age (range): Intervention, 75.9 (64–92)*  *Control, 74.9 (60–91)*  *Number of medications:* *Intervention:* NR  *Control:* NR  *HCPs:* 44 pharmacists | *Intervention type:* Patients had pharmaceutical care consultation with community pharmacist. Recommendations made to GP. The study pharmacist followed up with the patient clinically at 3, 6 and 12 months, updating of the pharmaceutical care plan as needed.  *Control group:* Wait-list control.  *Domains of integration:* Informational and clinical domains (moderate).  *Duration of intervention:* 12 months. | Primary  MAI  HRQoL, SF-36  Secondary  No. of inappropriate medications  Change in medications  Recommendation implementation by GP | *Mean MAI at 6-month follow-up:*  *Intervention*, 3.1  *Control,* 4.2  *Mean difference at 6 months*, 1.1 (95% CI -1.78 to -0.42, p=0.003)  *SF-36:*  *Emotional role:*  13.4-unit difference, P = 0.024  Favouring control  *Social functioning:*  7.7-unit difference, P = 0.019  Favouring control  Change in social functioning not clinically significant change |
| Campins (42) (2017, Spain) | *Total, n:* 503  *Age, mean (SD): Intervention,* 79.16 (5.5)  *Control,* 78.78 (5.46)  *Number of medications:* *Intervention:* 10.79 (2.52)  *Control:* 10.91 (2.65)  *HCPs: NR* | *Intervention type:* Pharmacist performed chart review. Pharmacist discussed recommendations with GP and therapeutic plan made. Recommendations were discussed with the patient, and a final decision was agreed by physicians and their patients in a face-to-face visit.  *Control group:* Usual GP care  *Domains of integration:* Organizational, informational, and clinical domains (moderate).  *Duration of intervention:* 12 months. | Primary  No. medications  Secondary  Medication appropriateness  Intervention Effectiveness  Change in medications  Adherence  (Morisky-Green)  HRQoL, EQ5D  Healthcare utilization  Mortality | *Mean number of medications at follow up:*  *Intervention*,10.03  *Control,* 10.91  P=0.001 |
| Geurts (43) (2016, The Netherlands) | *Total, n:* 512  *Age (mean, SD):* *Intervention:* 72.5 (7.735), *Control:* 73.1 (7.797)  *Number of medications:* *Intervention:* 8.3 (2.721)  *Control:* 7.9 (2.926)  *HCPs:* 8 pharmacies/primary care sites. | *Intervention type:* Community pharmacists had clinical medication review with patients. Recommendations made to GP. Implemented care intervention.  *Control group:* usual care.  *Domains of integration:* Organizational, informational, clinical, and normative domains (robust).  *Duration of intervention:* 18 months. | Primary  Decrease in potential DRPs and PCIs  Secondary  Biomarkers for: Blood pressure, dyslipidaemia, BMI, diabetes and renal function | *Total number of DRPs and PCIs at follow-up:*  *Intervention*, 208  *Control* not reported |
| Graffen (53) (2004, Australia) | *Total, n:* 402  *Age (median, range):* 77.7 (66-102)  *Number of medications (mean):* 8.4  *HCPs:* 8 general practices | *Intervention type:* Pharmacist reviewed medication profiles of rural patients in practice and made recommendations to GP. Changes were made by agreement between patient and GP.  *Control group:* usual care  *Domains of integration:* Organizational, informational, and clinical domains (moderate).  *Duration of intervention:* 18 months. | Primary  HRQoL SF-36  Secondary  Recommendation uptake  Hospitalizations | *SF-36*  Significant difference in vitality (p<0.009) and mental health (p<0.0001) in favour of intervention post-intervention. |
| Granas (39) (1998, UK) | *Total, n:* 285  *Age (median, range):* 65 (1 - 102)*:*  *Number of medications (median, range):* 5 (3-27)  *HCPs:*  1 community pharmacist  2 GPs | *Intervention type:* Pharmacist performed chart-based medication review in practice. Recommendations made to GP. GP followed up with patient.  *Control group:* Usual care.  *Domains of integration:* Organizational, informational, and clinical domains (moderate).  *Duration of intervention:* 24 months. | Primary  DRPs  Secondary  Recommendation uptake | *DRPs at follow-up*  *Intervention,* 7.8%  *Control,* 11.6%  Statistically significant difference (p<0.001).  Adjusted ARR, 26%. |
| Hanlon (34) (1996, USA) | *Total, n:* 208  *Age (mean, SD): Intervention*, 69.7 (3.5)  C*ontrol*, 69.9 (4.1)  *Number of medications (mean (*SD)*):*  *Intervention:* 7.6 (2.8)  *Control:* 8.2 (2.7)  *HCPs:* 2 pharmacists | *Intervention type:* Comprehensive medication review with the pharmacist in practice and follow-up at 11.5-13 months.  *Control group:* usual care with closeout interview by second pharmacist blinded to allocation.  *Domains of integration:* Organizational, informational, clinical, and financial domains (robust).  *Duration of intervention:* 13 months | Primary  Prescribing appropriateness; MAI  Secondary  Improvement in inappropriate prescribing  Medication compliance  Medication knowledge  Number of medications  HrQoL, SF-36 | *Adjusted MAI at 12 months (mean, SD)*  *Intervention,* 12.8 (0.7)  *Control,* 16.7 (0.7)  Inappropriate prescribing scores declined more in intervention than control at 12 months: 28% vs 5% (p=0.0002) |
| Jameson (35) (2001, USA) | *Total, n:* 168  *Age:*  *Intervention:* 51.4 (10.1) *Control:* 52.5 (10.6)  *Number of medications:* NR  *HCPs:* 1 pharmacist  133 physicians (in 4 practices) | *Intervention type:* Medication review with patient in practice. Discussed DRPs with GP. Care plan developed with GP. Changes discussed with patient.  *Control group:* Usual care.  *Domains of integration:* Organizational, informational, and clinical domains (moderate).  *Duration of intervention:* 9 months. | Primary  Medical/drug costs  Secondary  Adverse effects | *Medication cost at follow-up $*  *Intervention:* 1657 (1068)  *Control:* 1602 (1202)  No significant change. |
| Krska (40) (2001, UK) | *Total, n:* 381  *Age (mean, range):* *Intervention:* 74.8 (65-90), C*ontrol:* 75.2 (65-93)  *Number of medications (mean, SD):*  *Intervention:* 7.4±2.7  *Control:* 7.7±2.8  *HCPs:* NR | *Intervention type:* Pharmacist performed chart review followed by medication review with patient in their own home. Pharmaceutical care plan developed in practice. Notes forwarded to GP who indicated level of agreement. Implemented actions where appropriate.  *Control group:* Usual care.  *Domains of integration:* Organizational, informational, and clinical domains (moderate).  *Duration of intervention:* 3 months. | Primary  Pharmaceutical care issues  Secondary  Medication costs  HRQoL, SF-36  Healthcare utilization | *PCIs at 3 months*  *Intervention:* 256 (21.2%)  *Control:* 856 (60.7%)  *Crude ARR*: 39.5% |
| Kwint (44) (2011, The Netherlands) | *Total, n:* 118  *Age (mean* ± SD*):* *Intervention:* 78.7 (6.8) *Control:* 80.0 (7.2)  *Number of medications (mean* ± SD*):* *Intervention:* 10.3 (3.1)  *Control:* 9.8 (3.6)  *HCPs:* 6 community pharmacists; each community pharmacist recruited two GPs. | *Intervention type:* Community pharmacist collated information from pharmacy and GP. Data reviewed by 2 pharmacists. Results sent to community pharmacist to discuss with GP.  *Control group:* Wait-list control.  *Domains of integration:* Informational and clinical domains (moderate).  *Duration of intervention:* 6 months. | Primary  DRPs  Secondary  Medication changes  Recommendation’s uptake | *DRPs at baseline*  *Intervention:* 4.5  *Control:* 4.4  *DRPs at 6 months*  *Intervention:* 3.2  *Control:* 4.2 |
| Lenaghan (41) (2007, UK) | *Total, n:* 136  *Age:* 84.3  *Number of medications:* 9.45  *HCPs:* 1 community pharmacist  9 GPs based in one practice. | *Intervention type:* Comprehensive medication review in patient’s home. Community pharmacist had notes from both pharmacy and medical notes. Recommendations discussed and implemented with GP and discussed with patients. Further follow-up at 6-8 weeks.  *Control group:* Wait-list control.  *Domains of integration:* Organizational, informational, and clinical domains (moderate).  *Duration of intervention:* 6 months. | Primary  Unplanned hospital admissions  Secondary  Mortality  HRQoL, EQ5D  No. of medications | *Hospitalizations at follow-up:*  *Intervention,* 21  *Control,* 20  P = 0.8 |
| Sellors (36) (2003, Canada) | *Total, n:* 889  *Age (mean, SD):* *Intervention*: 74.0 (6.1)  *Control group:* 74.0 (6.0)  *Number of medications (mean):*  *Intervention:* 12.4  *Control:* 12.2  *HCPs:* n=24 pharmacists  n=48 physicians | *Intervention type:* Community pharmacists (who had additional post-university training in the prevention, identification, and resolution of drug-related problems) conducted face-to-face medication reviews with the patients and then gave written recommendations to the physicians to resolve any drug-related problems.  *Control group:* Usual care.  *Domains of integration:* Organizational, informational, clinical, and financial domains (robust).  *Duration of intervention:* 5 months. | Primary  DRPs  Secondary  Uptake of recommendations  Length of physician meeting  Reasons for not implementing recommendations  Cost  HRQoL, SF-36 | *Recommendations uptake at follow-up:*  Fully implemented: 46.3% (506/1093).  Partially implemented: 9.3% (102/1093).  Unsuccessful implementation: 16.7% (182/1093). |
| Taylor (37) (2003, USA) | *Total, n:* 69  *Age (mean, SD): Intervention:* 64.4 (13.7)  *Control:* 66.7 (12.3)  *Number of medications (mean, SD):*  *Intervention:* 6.3 (2.2)  *Control:* 5.7 (1.7)  *HCPs:* 4 pharmacists | *Intervention type:* Patients in the intervention group received usual care plus pharmacotherapeutic interventions by a pharmacist during regularly scheduled office visits before seeing a physician. Pharmacist developed education packs for patients with diabetes, hypertension, dyslipidaemia and anti-coagulation services.  *Control group:* Medical records review at baseline and 12 months later performed by pharmacist, no advice, or recommendations to patient/physician.  *Domains of integration:* Organizational, informational, clinical, and financial domains (robust).  *Duration of intervention:* 12 months. | Primary  Prescribing appropriateness (MAI) and ADRs  Secondary  Hospitalizations and ED visits  Hypertension outcomes  Biomarkers for: Diabetes, dyslipidaemia, blood pressure and anticoagulation services  HRQoL, SF-36 | *Inappropriate prescriptions at baseline (MAI)*  *Intervention:* 210  *Control:* 207  *Inappropriate prescriptions at follow-up*  *Intervention:* 155  *Control:* 224  Ratings for inappropriate prescribing improved in all 10 domains evaluated in the intervention group but worsened in 5 domains in the control group. |
| VanDerMeer (45) (2018, The Netherlands) | *Total, n:* 157  *Age (mean, SD): Intervention:* 75.7 (6.9)  C*ontrol:* 76.6 (6.7)  *Number of medications (mean, SD):*  *Intervention:* 8.4 (2.4)  *Control:* 9.3 (3.2)  *HCPs:* 15 community pharmacies | *Intervention type:* Pharmacotherapeutic review with community pharmacist. Written recommendations forwarded to GP followed by an MDT meeting where an action plan was decided. This was discussed with the patient and decisions taken were followed up.  *Control group:* Wait-list control.  *Domains of integration:* Organizational, informational, and clinical domains (moderate).  *Duration of intervention:* 3 months. | Primary  Change in DBI  Secondary  Presence of anticholinergic/sedative side effects  Falls  Cognitive function  Activities of daily living  HRQoL, EQ5D-3L  Hospital admission  Mortality | *Proportion of patients with a decrease of DBI≥0.5 at follow up*  *Intervention:* 17.3%  *Control:* 15.9%  OR 1.04, CI 0.47 to 2.64, p=0.927) |
| Verdoorn (46) (2019, The Netherlands) | *Total, n:* 629  *Age (range):*  *Intervention:* 80 (76-83)  *Control:* 78 (74-82)  *Number of medications (median, IQR):* 9.0 (7.5-10.5)  *HCPs:* 43 pharmacists  113 GPs | *Intervention type:* Patients received a clinical medical review from community pharmacist who had both pharmacy data and medical history. Potential DRPs reported and discussed face-to-face with GP. PCP proposed and actions agreed. Two follow-up appointments to review changes made.  *Control group:* Wait-list control.  *Domains of integration:* Organizational, informational, and clinical domains (moderate).  *Duration of intervention:* 6 months. | Primary  EQ5D-5L  EQ5D-VAS  No. of health problems  Secondary  Number of medications  Medications commenced or discontinued | *EQ5D-5L at follow up:*  Intervention, 0.73 (0.20)  Control, 0.74 (0.18)  *EQ5D-VAS at follow up:*  Intervention, 70 (16)  Control, 69 (15)  HR-QoL measured with EQ-VAS increased by 3.4 points (95% confidence interval [CI] 0.94 to 5.8; p = 0.006),  *Number of health problems with impact on daily life:* Decreased by 12% in intervention (difference at 6 months −0.34; 95% CI −0.62 to −0.044; p = 0.024) as compared with the control group. |
| Vinks (47) (2009, The Netherlands) | *Total, n:* 174  *Age (mean, SD):*  *Intervention:* 76.6 (6.5)  *Control:* 76.6 (6.4).  *Number of medications (mean, SD):*  *Intervention:* 8.8 (2.5)  *Control:* 8.5 (2.3)  *HCPs:* 16 pharmacies | *Intervention type:* Community pharmacists reviewed patients’ medications and compiled a list of recommended changes in medication was compiled by the pharmacist for the patients in the intervention group. Recommendations for medication change were discussed with the general practitioner (GP). Repeat screening conducted at follow-up.  *Control group:* Usual care.  *Domains of integration:* Informational and clinical domains (moderate).  *Duration of intervention:* 12 months. | Primary  DRPs  Secondary  No. of medications  Recommendations and uptake | *Mean DRPs at follow up*  *Intervention:* 3.29  *Control:* 3.62  Mean difference –16.3%; 95% CI –24.3, –8.3). |
| Zillich (38) (2014, USA) | *Total, n:* 961  *Age (mean, SD):* 73 (13)  *Number of medications:* *Intervention:* 14 (11)  *Control:* 13 (8)  *HCPs:* NR | *Intervention type:* Nurse completed admission procedures and forwarded meds info to MTM intervention provider. Pharmacy technician called to verify all meds. Community pharmacist then rang and completed comprehensive med review with patient/carer. Developed medication related action plan. Pharmacist provided follow-up phone call on day 7, and again at day 30 and 60 as needed.  *Control group:* Usual home health care.  *Domains of integration:* Organizational, informational, and clinical domains (moderate).  *Duration of intervention:* 60 days. | Primary  60-day hospitalizations  Secondary  MRPs  Recommendation uptake | *Hospitalizations at follow up:*  *Intervention:* 83  *Control:* 112  Adjusted OR: 1.26, 95 percent CI: 0.89–1.77, p=0.19). |
| **Cluster Randomised Controlled Trials** | | | | |
| Bernsten (2001) (48)  Multiple sites: (seven EU countries) | *Total, n:* 2454  *Age (median, IQR):* 74(8)  *Number of medications:* *Intervention:* 7.05 (2.51)  *Control:* 6.97 (2.51)  *HCPs:* 190 community pharmacies. | *Intervention type:* Pharmacists completed study training and informed local GPs and formed formal links. Patient received pharmaceutical care intervention in collaboration with GPs.  *Control group:* Usual care.  *Domains of integration:* Clinical domain (minimum).  *Duration of intervention:* 18 months. | Primary  HRQoL; SF-36  Secondary:  Hospitalizations  Cost  Compliance  Recommendation acceptance | *HRQoL:*  A general decline in health-related quality of life over time was observed in the pooled data; however, improvements were achieved in patients involved in the pharmaceutical care programme in some countries. |
| Sorensen (54) (2004, Australia) | *Total, n:* 400  *Age (mean, range):* Intervention 72.3 (37-100), control 71.4 (25-99)  *Number of medications:* *Intervention:* 9.7 (9.1-10.3)  *Control:* 8.9 (8.3-9.4)  *HCPs:* n=53 pharmacists  n=92 GPs | *Intervention type:* GPs were the units of randomization. GPs made referrals to the community pharmacist who conducted medication review based on pharmacy data and medical records. Prepared report for GP, recommendations discussed at MDT meeting and action plan developed. GP implemented plan with patient agreement.  *Control group:* Usual care.  *Domains of integration:* Organizational and clinical domains (moderate).  *Duration of intervention:* 6 months. | Primary  DUSOI-A  Secondary  Problems and recommendations  HRQoL, SF-36  ADEs  Costs | *Change in DUSOI-A at follow up:*  Intervention, reduced by 4.92  Control, reduced by 1.34 |
| Varas-Doval (49) (2020, Spain) | *Total, n:* 1403  *Age (mean, SD):*  *Intervention:* 75.34 (6.46)  *Control:* 74.92 (6.59)  *Number of medications (mean, SD):*  *Intervention:* 7.74 (2.5)  *Control:* 7.39 (2.37)  *HCPs:* n=178 pharmacies, n=250 pharmacists. | *Intervention type:* Community pharmacists provided Medication review with follow up (MRF). Pharmacist communicated with GPs via face to face or telephone. Follow up on a monthly basis for duration of the intervention.  *Control group:* Usual care.  *Domains of integration:* Organizational and clinical domains (moderate).  *Duration of intervention:* 8 months | Primary  Uncontrolled health problems  Secondary  DRPs  Interventions made by pharmacists | *Uncontrolled health problems at follow up [mean (95% CI)]:*  *Intervention:* 0.65 (0.43, 0.88)  *Control:* 0.69 (0.47, 0.91)  *Reduction in the number of uncontrolled health problems:*  *Intervention:* -0.72 (95% CI: -0.80, -0.65)  *Control:* -0.03 (95%CI: -0.10, 0.04) |
| **Non-Randomised Controlled Trials** | | | | |
| Leendertse (50) (2013, The Netherlands) | *Total, n:* 674  *Age (95% CI) :*  *Intervention:* 75.8* (74.9–76.4)  *Control:* 75.7* (75.1–76.7).  *Number of medications:* *Intervention:* 7.8* (7.7- 8.2)  *Control:* 7.9* (7.5 – 8.2)  *HCPs:* 42 primary health care settings. 1 intervention and 1 control GP. Number of GPs and pharmacists not reported.  * Values are results of the linear mixed-effects model. | *Intervention type:* In each practice, patients were recruited from community pharmacy, Community pharmacist conducted structured pharmacotherapeutic review with patient based on pharmacy history and medical notes. Pharmacist and GP met to discuss PCP. Agreed changes implemented and monitored by GP/practice nurse.  *Control group:* Usual care.  *Domains of integration:* Organizational, informational, and clinical domains (moderate).  *Duration of intervention:* 12 months. | Primary  Medication-related hospitalizations  Secondary  Survival  HRQoL, EQ5D  ADEs  Drug therapy problems and care issues.  Interventions recommended | *Hospitalizations at follow-up:*  *Intervention:* 6  *Control:* 10  Reduction in hospitalizations in intervention 1.6% vs 3.2% (HR 0.50, 95%CI 0.12-1.59). |
| **Controlled Before-After** | | | | |
| Sloeserwij (51) (2019, The Netherlands) | *Total, n:* 11928  *Age (mean, SD): 75 (8)*  *Number of medications:* 6 (5-8)  *HCPs:* 9 pharmacists  25 general practices | *Intervention type:* Pharmacists embedded in general practices for three months prior to intervention. Pharmaceutical care for high-risk patients- pharmacist performed medication reviews with patients and medications reconciliation amongst other practice-related activities.  *Control group:* Usual care was normal GP review. Usual care plus was medication review conducted by accredited community pharmacist.  *Domains of integration:* Organizational, informational, and clinical domains (moderate).  *Duration of intervention:* 12 months. | Primary  Medication-related hospitalizations  Secondary  DBI  Costs | *Medication-related hospitalizations at follow up:*  *Intervention:* 230  *Usual care:* 355  *Usual care plus:* 237  Rate ratio of medication‐related hospitalizations in the intervention group compared to usual care was 0.68 (95% CI: 0.57–0.82) and 1.05 (95% CI: 0.73–1.52) compared to usual care plus. |

Text highlighted in bold indicate main column headings. Text highlighted in italics are subheadings located within a column.

Key; HCP (healthcare professional) RCT (randomised controlled trial), CBA (controlled before-after trial), HRQoL (health related quality of life), SF-36 (short form 36), MAI (medications appropriateness index), EQ5D (EuroQoL 5 domains), HbA1c (haemoglobin A1C), LDL (low-density lipoprotein), DRP (drug related problem), ATC (Anatomical Therapeutic Chemical), PCI (pharmaceutical care issue), ARR (absolute risk reduction), NNT (number needed to treat), NR, not reported; HR (hazard ratio), CI (confidence interval), ITS (interrupted time series), PCT (primary care trust) DBI (drug burden index), MDT (multi-disciplinary team), DUSOI-A (Duke’s Severity of Illness Visual Analogue Scale), MRP (medication-related problem), VAS (visual analogue scale), MTM (medications therapeutic management), PDTP (potential drug therapy problem), CBA (controlled before-after)
