## Supplementary material for "The effectiveness and cost of integrating pharmacists within general practice to optimize prescribing and health outcomes in primary care patients with polypharmacy: A systematic review": Table 3

### Table 3 Characteristics of included economic studies

| **Author (year, study)** | **Country perspective** | **Analysis type** | **Follow-up/time horizon** | **Control** | **Intervention** | **Results** |
| --- | --- | --- | --- | --- | --- | --- |
| **Campins (2019, Campins study)** | Third party payer, public, Spain | CBA | 12 months post-intervention | Usual GP care | Pharmacist performed chart review. Pharmacist discussed recommendations with GP and therapeutic plan made. Recommendations were discussed with the patient, and a final decision was agreed by physicians and their patients in a face-to-face visit. | Drug expenditure savings per patient/year attributable to intervention: €88.42  Drug costs reduction: Intervention: €321.43 (CI=233.77–409.79) Control: €232.94 (CI=141.64–323.15) Intervention cost: €37.17  The estimated return per Euro invested in the programme: €3.27 per patient a year on average. |
| **Cowper (1998, Hanlon study)** | Third party payer, public, US | CEA | 12 months | Usual care with closeout interview by second pharmacist blinded to allocation. | Comprehensive medication review with the pharmacist in practice and follow-up at 11.5-13 months. | Cost-effectiveness as reported by MAI score:  €1122.64/one unit change (estimated by researchers) |
| **Jodar-Sanchez (2015, Varas-Doval study)** | Third party payer, public, Spain | CUA | 12 months | Usual care | Community pharmacists provided Medication review with follow up (MRF). Pharmacist communicated with GPs via face to face or telephone. Follow up on a monthly basis for duration of the intervention. | Incremental effectiveness (intervention vs control)  0.0156 QALYs (SD = 0.004) (95% CI 0.008– 0.023)  Mean incremental total cost of intervention  -€321.88 ± 190.95 (95 % CI -696.14 to 52.37) |
| **Noain (2017, Varas-Doval study)** | Third party payer, public, Spain | Micro-costing, cost analysis | 12 months | Usual care | Community pharmacists provided medication review to patients with follow-up with GPs and patients. Same study as Jodar-Sanchez above. | Service provider cost per patient ranged from €251.72 (SD 90.5) to €398.12 (SD 164.4).  The mean initial investment per pharmacy was €5899.92 and the mean annual maintenance costs €3940.13  The potential service price ranged from €304.37 to €806.52 per patient a year. |
| **Malone (2000, Carter study)** | Third party payer, public, US | Cost analysis | Not reported, costs calculated for 12 months prior to trial and 12 months of trial. | Usual GP care | Patients were assessed by pharmacists at least three times (baseline, 6 & 12 months) but could see the patients as often as was medically indicated. Documentation via a standardised form was used for all patient encounters. Pharmacists saw patients in clinic or spoke on the phone. Discussed recommendations with GP. | Mean increases in total health care costs were €1553.89 for the intervention group and €2000.25 for the control group (p=0.06) |
| **Effectiveness studies with cost and cost-effectiveness data** | | | | | | |
| **Bernsten (2001)** | Third party payer, pan-European study, reported from Sweden | Cost analysis | 18-month trial period | Usual care | Pharmacists completed study training and informed local GPs and formed formal links. Patient received pharmaceutical care intervention in collaboration with GPs. | *Total cost:*  Between-group analysis indicated that there were no significant differences between the total cost for control and intervention patients in any country (Mann-Whitney, p > 0.05)  *Hospitalisations*  Germany, intervention patients had significantly lower costs associated with hospitalisations and contacts with specialists compared with control patients in the second 6-month period (Mann-Whitney test, p < 0.05)  *Cost savings*  Cost savings were achieved in most of the countries at each assessment period. |
| **Britton (1991)** | Third party payer, US | Cost analysis | 3-month trial period | Usual care | Pharmacist reviewed patient treatment and cost. Medication profile review form attached to each patient file for review by GP. Post GP consult, the pharmacist reviewed files for no. meds, meds changes and compliance. | *Intervention total cost savings:* €287.93  *Control total cost savings:* costs increased by €1295.93  *Intervention total cost avoidance:* €1588.39 |
| **Jameson (2001)** | Third party payer, US | Cost analysis | 9-month trial period | Usual care | Medication review with patient in practice. Discussed DRPs with GP. Care plan developed with GP. Changes discussed with patient. | No significant differences were demonstrated in the changes in medical or drug costs between the consult and the control groups  *Drug Changes at follow-up*  *Intervention:*  Change €73.87 (904.06)  *Control:*  Change €23.45 (11257.98)  *Medical changes at follow-up*  *Intervention:*  Change €632.02 (11716.46)  *Control:*  Change €367.02 (12231.22) |
| **Krska (2001)** | Third party payer, UK | Cost analysis | 3-month trial period | Usual care | Pharmacist performed chart review followed by medication review with patient in their own home. Pharmaceutical care plan developed in practice. Notes forwarded to GP who indicated level of agreement. Implemented actions where appropriate. | No significant difference at baseline or follow-up.  *Intervention Medication Costs (mean, SD)*  Baseline: €76.27 (56.52)  Follow-up: €75.49 (57.55)  *Control Medication Costs*  Baseline: €83.21 (65.13)  Follow-up: €82.84 (61.90) |
| **Sellors (2003)** | Third party payer, Canada | Cost analysis | 5-month trial period | Usual care | Community pharmacists (who had additional post-university training in the prevention, identification, and resolution of drug-related problems) conducted face-to-face medication reviews with the patients and then gave written recommendations to the physicians to resolve any drug-related problems. | *Mean daily cost of medications per patient (intervention vs control)*  €4.10 vs €3.94 (p = 0.72)  *Mean total cost of medications per patient (intervention vs control)*  €2.92 vs €3.08 (p = 0.78)  *Mean cost of healthcare resources per patient (intervention vs control)*  €1047.97 vs €1062.78 (p = 0.45) |
| **Sloeserwij (2019)** | Third party payer, Netherlands | Cost analysis | 12-month trial period | Usual care was normal GP review. Usual care plus was medication review conducted by accredited community pharmacist. | Pharmacists embedded in general practices for three months prior to intervention. Pharmaceutical care for high-risk patients- pharmacist performed medication reviews with patients and medications reconciliation amongst other practice-related activities. | Intervention vs usual care only. No difference in healthcare or medication costs reported.  *Ratio of healthcare costs in intervention group vs usual care group (95% CI)*  Primary care costs: 1.08 (0.99–1.17) p=0.073  Secondary care costs: 0.92 (0.65–1.29) p=0.622  Medication costs: 1.04 (0.98–1.10) p=0.172  *Secondary healthcare costs related to hospitalisations:*  No differences: adjusted ratio 0.82 (95% CI 0.64–1.06). |
| **Sorensen (2004)** | Third party payer, Australia | CEA | 6-month trial period | Usual care | GPs were the units of randomization. GPs made referrals to the community pharmacist who conducted medication review based on pharmacy data and medical records. Prepared report for GP, recommendations discussed at MDT meeting and action plan developed. GP implemented plan with patient agreement. | The cost–effectiveness ratio for the intervention based on cost savings, reduced adverse events and improved health outcomes was small.  *Cumulative cost/patient over the 8 months from enrolment*  Intervention: €5806.45  Control: €6160.15  *Net cost saving per intervention patient (marginal cost benefit*) was €58.05 per patient relative to controls.  *Incremental cost–effectiveness ratio in reducing ADEs and in improving DUSOI-A for the groups*  ADEs: €74.18  DUSOI-A: €69.88 |

Text highlighted in bold indicate main column headings. Text highlighted in italics are subheadings located within a column.

Key: CBA (cost benefit analysis), CI (confidence interval), CEA (cost-effectiveness analysis), CUA (cost utility analysis), MRF (medication review with follow up), MAI (medications appropriateness index), QALY (quality adjusted life years), SD (standard deviation), DRPs (drug related problems), ADEs (adverse drug events), DUSOI-A (Duke’s Severity of Illness Visual Analogue Scale)
