## Supplementary material for "The effectiveness and cost of integrating pharmacists within general practice to optimize prescribing and health outcomes in primary care patients with polypharmacy: A systematic review": Table 4

### Table 4 Summary of Findings table

| **Title** The effectiveness and cost of integrating pharmacists within general practice to optimize prescribing and health outcomes in primary care patients with polypharmacy | | | |
| --- | --- | --- | --- |
| **Patients or population:** Patients over the age of 65 on ten or more medications  **Settings:** Primary care  **Intervention:** Pharmacist integration to optimise medications and improve patient outcomes  **Comparison:** Usual care | | | |
| **Outcomes** | **Impact** | **Number of  participants (Studies)** | **Certainty of the evidence (GRADE)** |
| **Potentially inappropriate prescribing** | Ten studies favoured pharmacist integration, eight of which demonstrated signficant changes in favour of the pharmacist integration group. | 1486 participants (10 studies) | ⊕⊕⊕⊖^a^ Moderate |
| **Number of medications** | Mean difference -0.80 [-1.17, -0.43]. Direction of effect of four of the seven studies favoured pharmacist integration in reducing the number of medications prescribed. Confidence intervals for three studies included zero. | 1176 participants (7 studies) | ⊕⊕⊕⊖^a^ Moderate |
| **Health-related quality of life** | Unclear effect, the direction of results could not be determined due to the heterogeneity in reported results. | 4535 participants (15 studies) | ⊕⊖⊖⊖^a, b, c^ Very low |
| **Adverse drug events** | Unclear effect, pharmacist integration tended to reduce the risk of ADEs, two studies reported significant results and two studies did not. | 409 participants (4 studies) | ⊕⊕⊖⊖^a, c^ Low |
| **Mortality** | No clear effect on mortality. | 327 participants (2 studies) | ⊕⊕⊖⊖^d^ Low |
| GRADE Working Group grades of evidence  **High** = This research provides a very good indication of the likely effect. The likelihood that the effect will be substantially different is low.  **Moderate** = This research provides a good indication of the likely effect. The likelihood that the effect will be substantially different is moderate.  **Low** = This research provides some indication of the likely effect. However, the likelihood that it will be substantially different is high.  **Very low** = This research does not provide a reliable indication of the likely effect. The likelihood that the effect will be substantially different is very high. | | | |
| Footnotes:  a; downgrade by one level due to serious concerns relating to risk of bias  b; downgrade by one level due to serious concerns relating to inconsistency of results  c; downgrade by one level due to serious concerns relating to imprecision of results  d; downgrade by two levels due to very serious concerns relating to imprecision of results | | | |

Text highlighted in bold indicate main headings.
