## Supplementary material for "The effectiveness and cost of integrating pharmacists within general practice to optimize prescribing and health outcomes in primary care patients with polypharmacy: A systematic review": Figure 1

Records identified from database searching

(Date of inception-January 2021, n= 26887)

Records screened

(n = 16882)

Records excluded

(n = 16675)

Full texts assessed for eligibility (n = 207)

Full texts excluded (n = 179):

Conference abstract (n = 38)

Single condition focus (n = 40)

Based on PICO (n=62)

Wrong study design (n = 16)

Protocol paper (n = 7)

Study ongoing: no results available (n = 7)

Duplicate paper (n = 3)

Excluded following discussion between reviewers (n = 6) Reasons for exclusion; wrong study design, single condition focus, less than 80% of patients with polypharmacy

Full texts included in review

28 papers reporting 23 studies included in the narrative synthesis

7 studies included in meta-analysis

**Identification**

**Screening**

**Included**

Records after duplicates removed (n = 19035)

Records screened for clearly ineligible reports (n = 2153)

**Eligibility**
