## Supplementary figures and images for "The effectiveness and cost of integrating pharmacists within general practice to optimize prescribing and health outcomes in primary care patients with polypharmacy: A systematic review"

### Figure 2

Figure 2

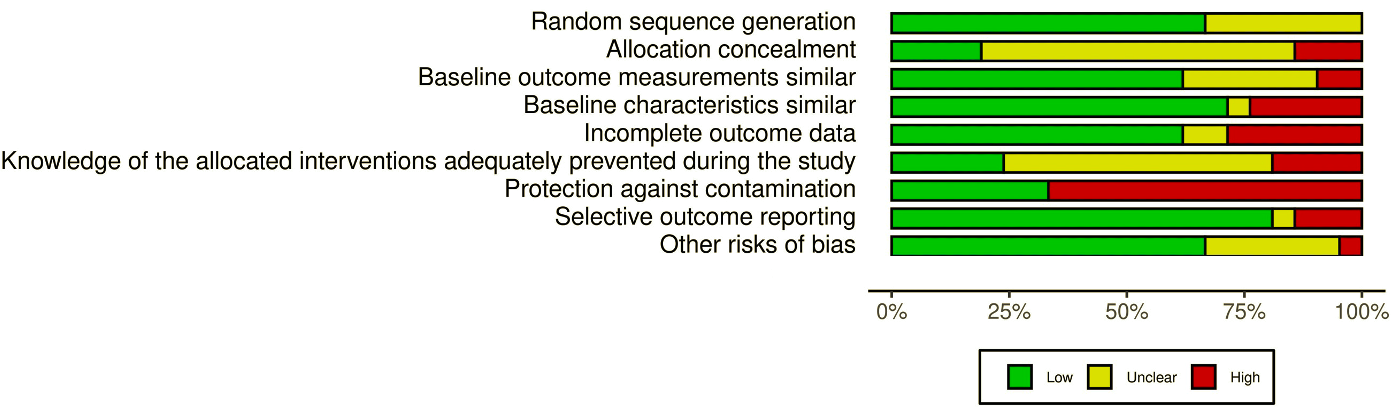

### Figure 3

Figure 3

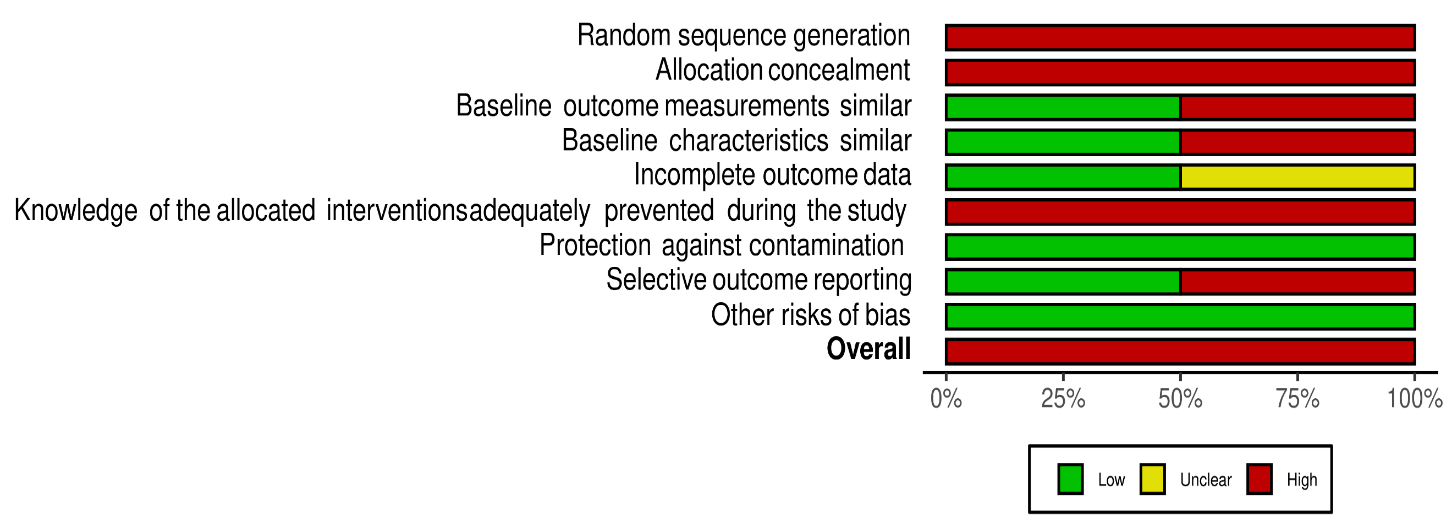

### Figure 4

Figure 4

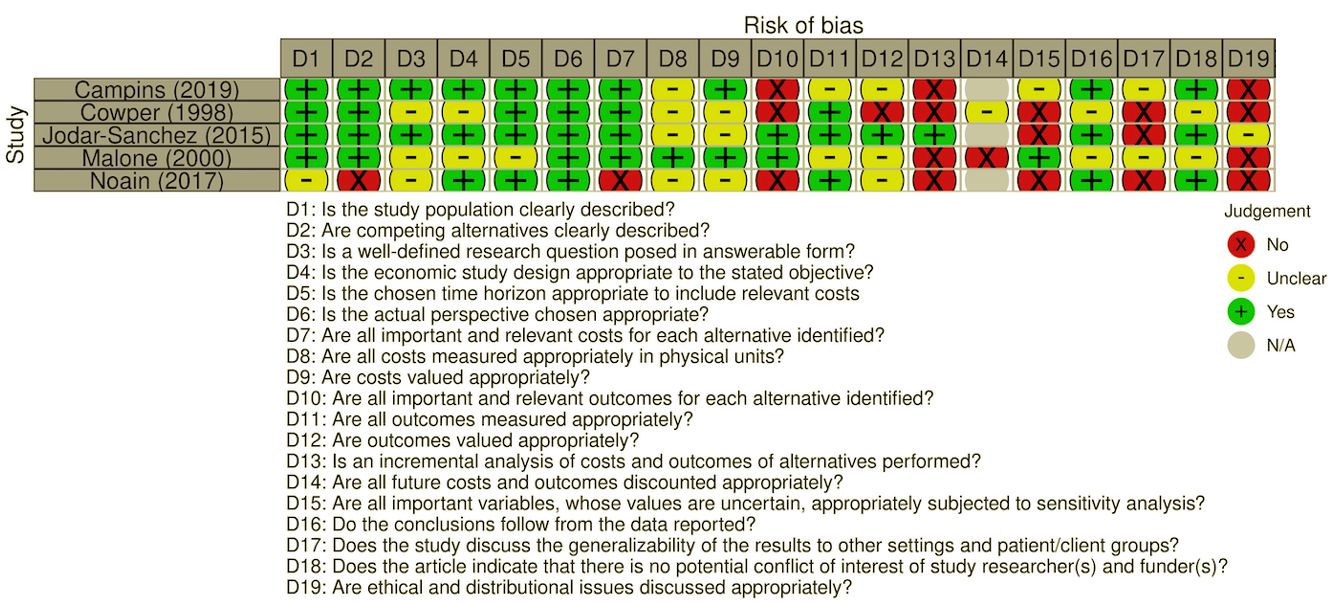

### Figure 5

Figure 5

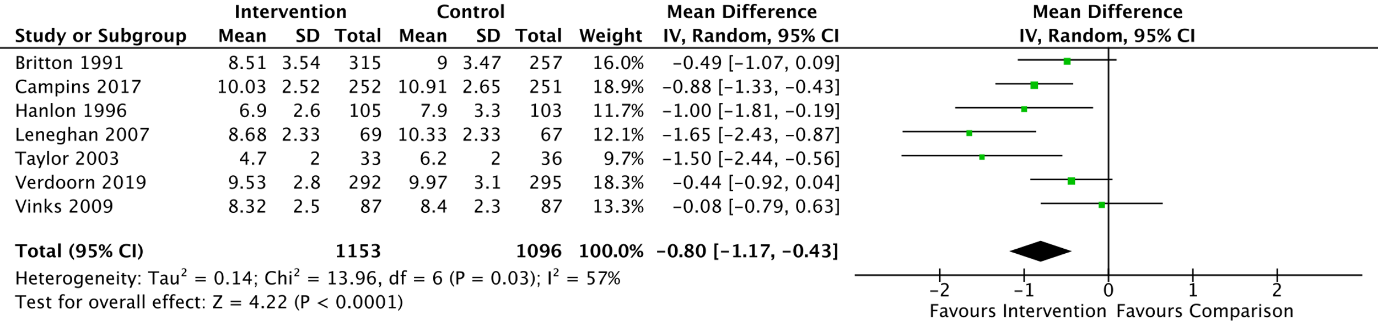
