## Additional File 1 for "The effectiveness and cost of integrating pharmacists within general practice to optimize prescribing and health outcomes in primary care patients with polypharmacy: A systematic review"

### PubMed Search Strategy

| Query | Items found |
| --- | --- |
| Search ((((((((("General Practice"[Mesh] OR "Primary Health Care"[Mesh]) OR "Community Health Services"[Mesh]) OR ("General Practitioners"[Mesh] OR "Physicians, Family"[Mesh])) OR ("Community Health Centers"[Mesh] OR "Physicians' Offices"[Mesh])) OR ((((general practice[Title/Abstract] OR family practice[Title/Abstract]) OR primary health care[Title/Abstract]) OR primary healthcare[Title/Abstract]) OR community health[Title/Abstract])) OR (((family practitioner[Title/Abstract] OR family practitioners[Title/Abstract]) OR (general practitioner[Title/Abstract] OR general practitioner's[Title/Abstract] OR general practitioners[Title/Abstract] OR general practitioners,[Title/Abstract])) OR (family physician[Title/Abstract] OR family physicians[Title/Abstract]))) AND (((("Pharmacies"[Mesh] OR "Pharmacy"[Mesh]) OR "Pharmaceutical Services"[Mesh]) OR "Pharmacists"[Mesh]) OR ((pharmacy[Title/Abstract] OR pharmacies[Title/Abstract]) OR pharmaceutical[Title/Abstract]))))) AND (((((((((((((randomized controlled trial[Publication Type]) OR controlled clinical trial[Publication Type]) OR pragmatic clinical trial[Publication Type]) OR multicenter study[Publication Type]) OR "Non-Randomized Controlled Trials as Topic"[Mesh]) OR "Interrupted Time Series Analysis"[Mesh]) OR "Controlled Before-After Studies"[Mesh]) OR (((randomis*[Title/Abstract]) OR randomiz*[Title/Abstract]) OR randomly[Title/Abstract])) OR groups[Title/Abstract]) OR (((((trial[Title]) OR multicenter[Title]) OR multi center[Title]) OR multicentre[Title]) OR multi centre[Title])) OR (((((((((intervention*[Title/Abstract]) OR effect*[Title/Abstract]) OR impact*[Title/Abstract]) OR controlled[Title/Abstract]) OR control group[Title/Abstract])) OR ((before[Title/Abstract]) AND after[Title/Abstract])) OR ((((pretest[Title/Abstract]) OR pre test[Title/Abstract])) AND ((posttest[Title/Abstract]) OR post test[Title/Abstract]))) OR (((((((quasiexperiment*[Title/Abstract]) OR quasi experiment*[Title/Abstract]) OR evaluat*[Title/Abstract]) OR time series[Title/Abstract]) OR time point[Title/Abstract]) OR time points[Title/Abstract]) OR repeated measur*[Title/Abstract])))) NOT (((((("Animals"[Mesh]) NOT (("Animals"[Mesh]) AND "Humans"[Mesh]))) OR (((((review[Publication Type]) OR meta analysis[Publication Type]) OR news[Publication Type]) OR comment[Publication Type]) OR editorial[Publication Type])) OR "The Cochrane database of systematic reviews"[Journal]) OR ((systematic review[Title]) OR literature review[Title]))) | 8209 |
| Search ((((((((((((randomized controlled trial[Publication Type]) OR controlled clinical trial[Publication Type]) OR pragmatic clinical trial[Publication Type]) OR multicenter study[Publication Type]) OR "Non-Randomized Controlled Trials as Topic"[Mesh]) OR "Interrupted Time Series Analysis"[Mesh]) OR "Controlled Before-After Studies"[Mesh]) OR (((randomis*[Title/Abstract]) OR randomiz*[Title/Abstract]) OR randomly[Title/Abstract])) OR groups[Title/Abstract]) OR (((((trial[Title]) OR multicenter[Title]) OR multi center[Title]) OR multicentre[Title]) OR multi centre[Title])) OR (((((((((intervention*[Title/Abstract]) OR effect*[Title/Abstract]) OR impact*[Title/Abstract]) OR controlled[Title/Abstract]) OR control group[Title/Abstract])) OR ((before[Title/Abstract]) AND after[Title/Abstract])) OR ((((pretest[Title/Abstract]) OR pre test[Title/Abstract])) AND ((posttest[Title/Abstract]) OR post test[Title/Abstract]))) OR (((((((quasiexperiment*[Title/Abstract]) OR quasi experiment*[Title/Abstract]) OR evaluat*[Title/Abstract]) OR time series[Title/Abstract]) OR time point[Title/Abstract]) OR time points[Title/Abstract]) OR repeated measur*[Title/Abstract])))) NOT (((((("Animals"[Mesh]) NOT (("Animals"[Mesh]) AND "Humans"[Mesh]))) OR (((((review[Publication Type]) OR meta analysis[Publication Type]) OR news[Publication Type]) OR comment[Publication Type]) OR editorial[Publication Type])) OR "The Cochrane database of systematic reviews"[Journal]) OR ((systematic review[Title]) OR literature review[Title])) | 7701502 |
| Search ((((((("General Practice"[Mesh] OR "Primary Health Care"[Mesh]) OR "Community Health Services"[Mesh]) OR ("General Practitioners"[Mesh] OR "Physicians, Family"[Mesh])) OR ("Community Health Centers"[Mesh] OR "Physicians' Offices"[Mesh])) OR ((((general practice[Title/Abstract] OR family practice[Title/Abstract]) OR primary health care[Title/Abstract]) OR primary healthcare[Title/Abstract]) OR community health[Title/Abstract])) OR (((family practitioner[Title/Abstract] OR family practitioners[Title/Abstract]) OR (general practitioner[Title/Abstract] OR general practitioner's[Title/Abstract] OR general practitioners[Title/Abstract] OR general practitioners,[Title/Abstract])) OR (family physician[Title/Abstract] OR family physicians[Title/Abstract]))) AND (((("Pharmacies"[Mesh] OR "Pharmacy"[Mesh]) OR "Pharmaceutical Services"[Mesh]) OR "Pharmacists"[Mesh]) OR ((pharmacy[Title/Abstract] OR pharmacies[Title/Abstract]) OR pharmaceutical[Title/Abstract]))) | 16510 |
