## Additional File 2 for "The effectiveness and cost of integrating pharmacists within general practice to optimize prescribing and health outcomes in primary care patients with polypharmacy: A systematic review"

### Domains of integration

| **Study** | **Organisational** | **Informational** | **Clinical** | **Functional** | **Normative** | **Financial** |
| --- | --- | --- | --- | --- | --- | --- |
| **Carter** | ✓ | ✓ | ✓ |  |  | ✓ |
| **Hanlon** | ✓ | ✓ | ✓ |  |  | ✓ |
| **Sellors** | ✓ | ✓ | ✓ |  |  | ✓ |
| **Taylor** | ✓ | ✓ | ✓ |  |  | ✓ |
| **Geurts** | ✓ | ✓ | ✓ |  | ✓ |  |
| **Britton** | ✓ | ✓ | ✓ |  |  |  |
| **Campins** | ✓ | ✓ | ✓ |  |  |  |
| **Graffen** | ✓ | ✓ | ✓ |  |  |  |
| **Granas** | ✓ | ✓ | ✓ |  |  |  |
| **Jameson** | ✓ | ✓ | ✓ |  |  |  |
| **Krska** | ✓ | ✓ | ✓ |  |  |  |
| **Leendertse** | ✓ | ✓ | ✓ |  |  |  |
| **Lenaghan** | ✓ | ✓ | ✓ |  |  |  |
| **Sloeserwij** | ✓ | ✓ | ✓ |  |  |  |
| **Van der Meer** | ✓ | ✓ | ✓ |  |  |  |
| **Verdoorn** | ✓ | ✓ | ✓ |  |  |  |
| **Zillich** | ✓ | ✓ | ✓ |  |  |  |
| **Sorensen** | ✓ |  | ✓ | ✓ |  |  |
| **Bryant** |  | ✓ | ✓ |  |  |  |
| **Kwint** |  | ✓ | ✓ |  |  |  |
| **Vinks** |  | ✓ | ✓ |  |  |  |
| **Varas-Doval** | ✓ |  | ✓ |  |  |  |
| **Bernsten** |  |  | ✓ |  |  |  |

Text highlighted in bold indicates a heading.
