## Additional File 3 for "The effectiveness and cost of integrating pharmacists within general practice to optimize prescribing and health outcomes in primary care patients with polypharmacy: A systematic review"

### 1. RoB assessment for PIP

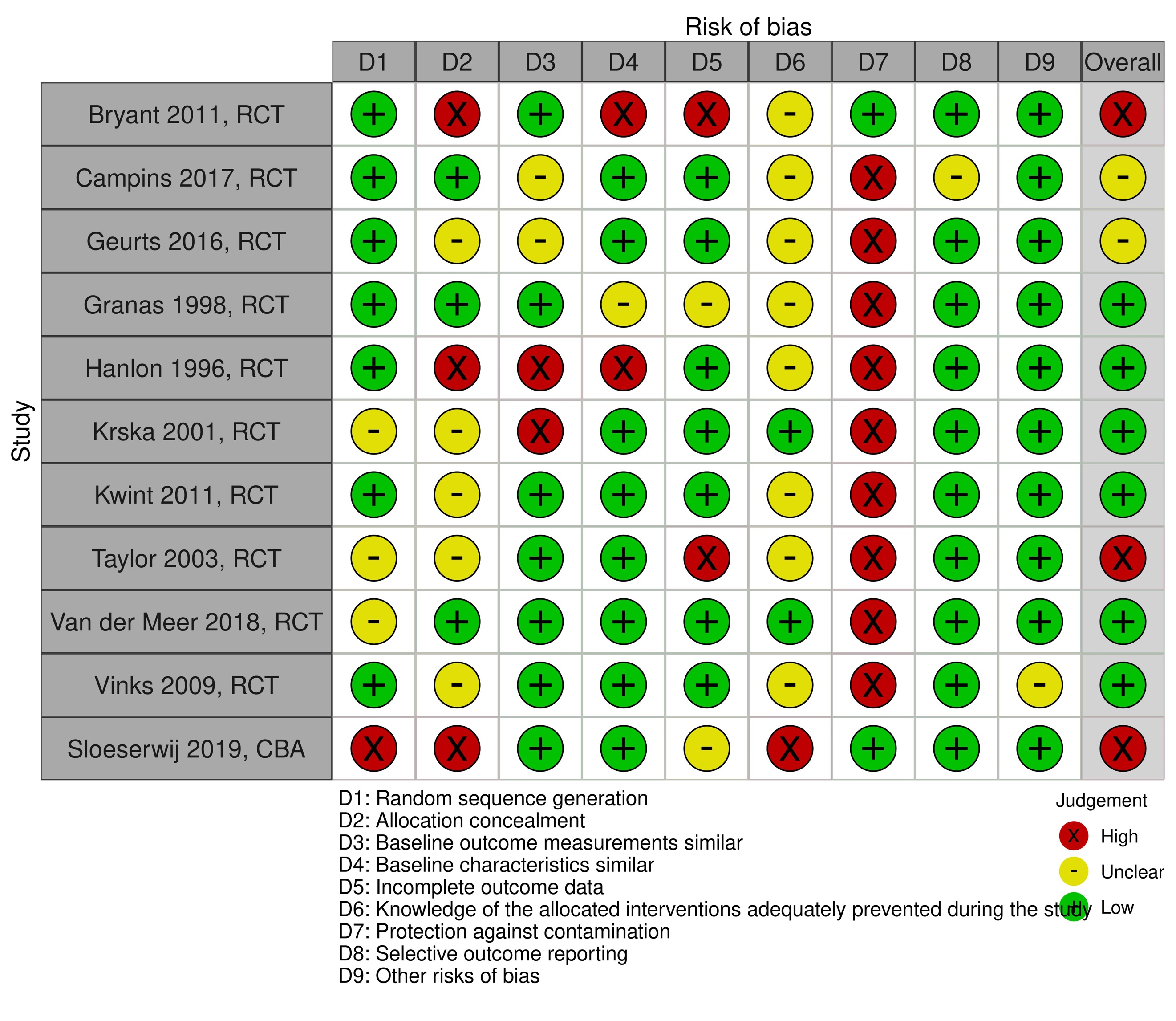

### 2. RoB assessment for number of medications

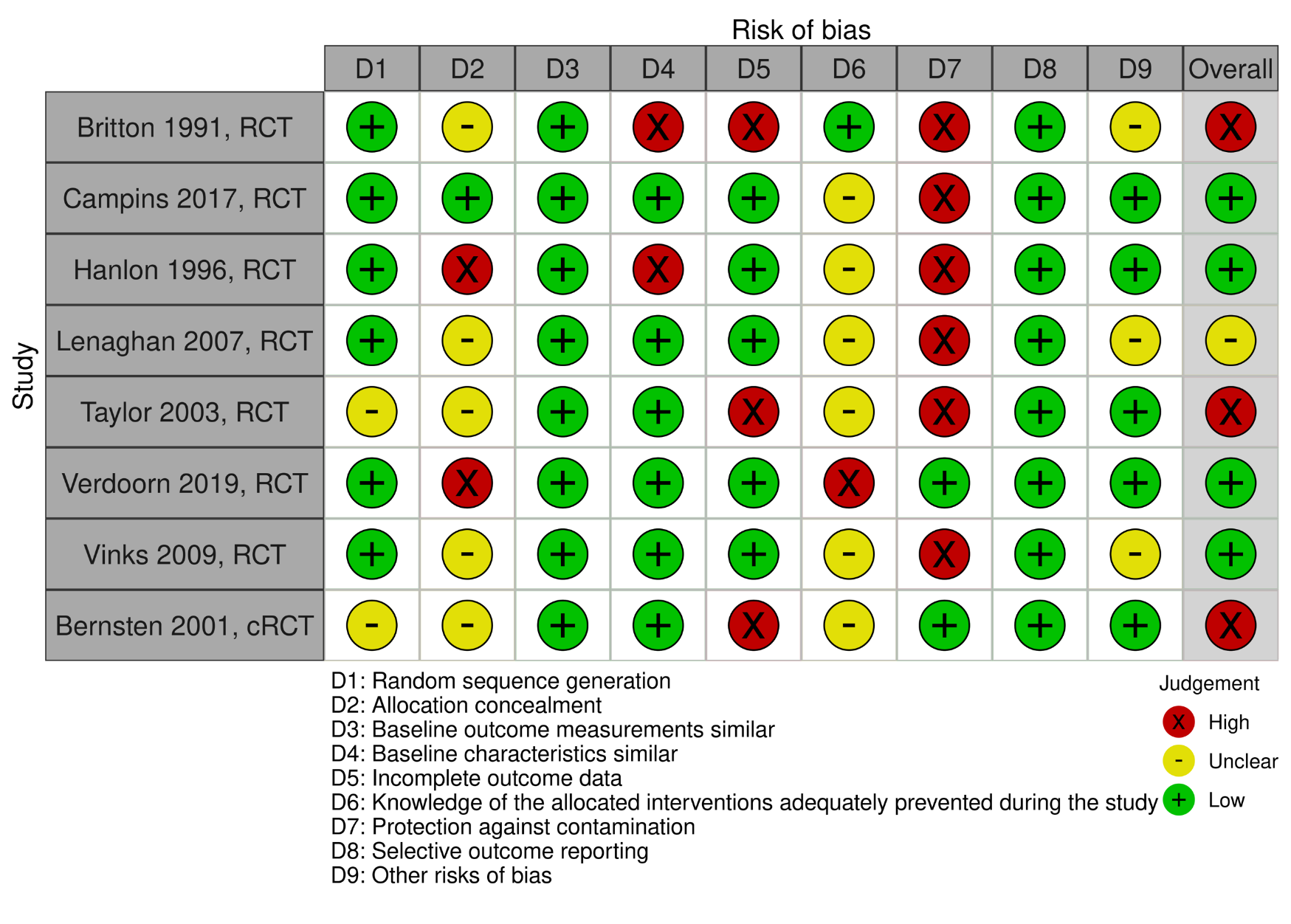

### 3. RoB assessment for HRQoL

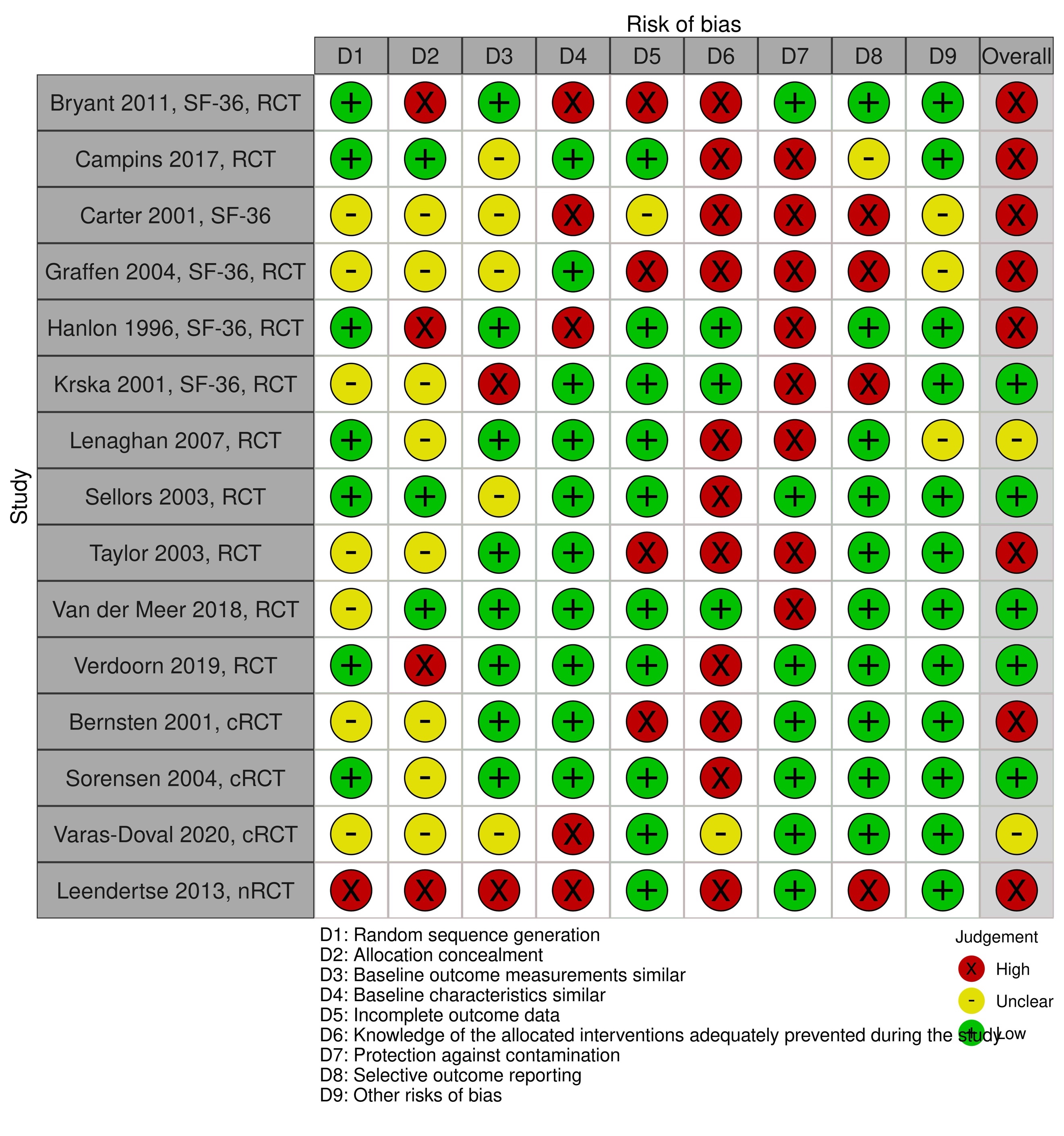

### 4. RoB ADRs

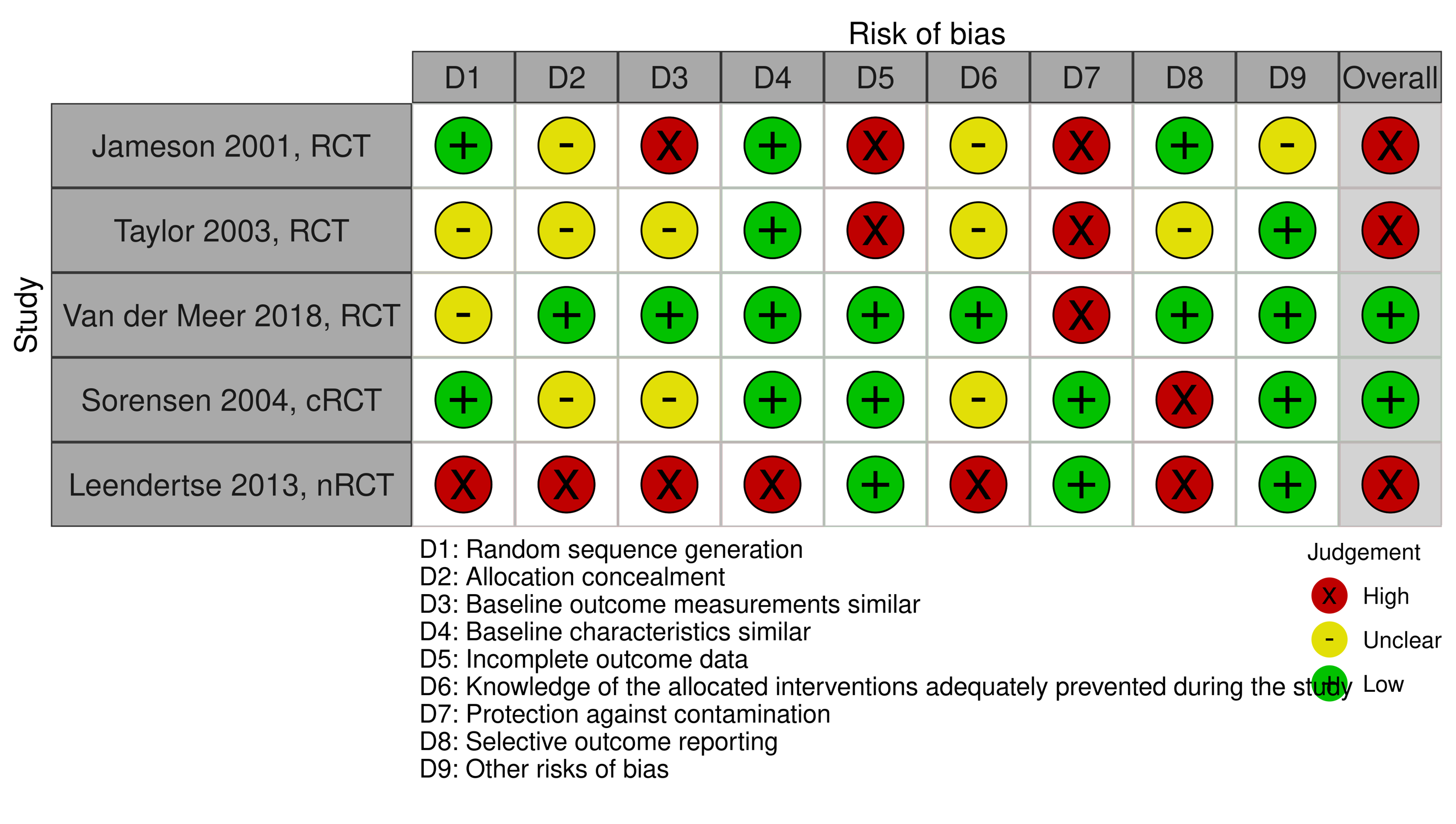

### 5. RoB assessment mortality

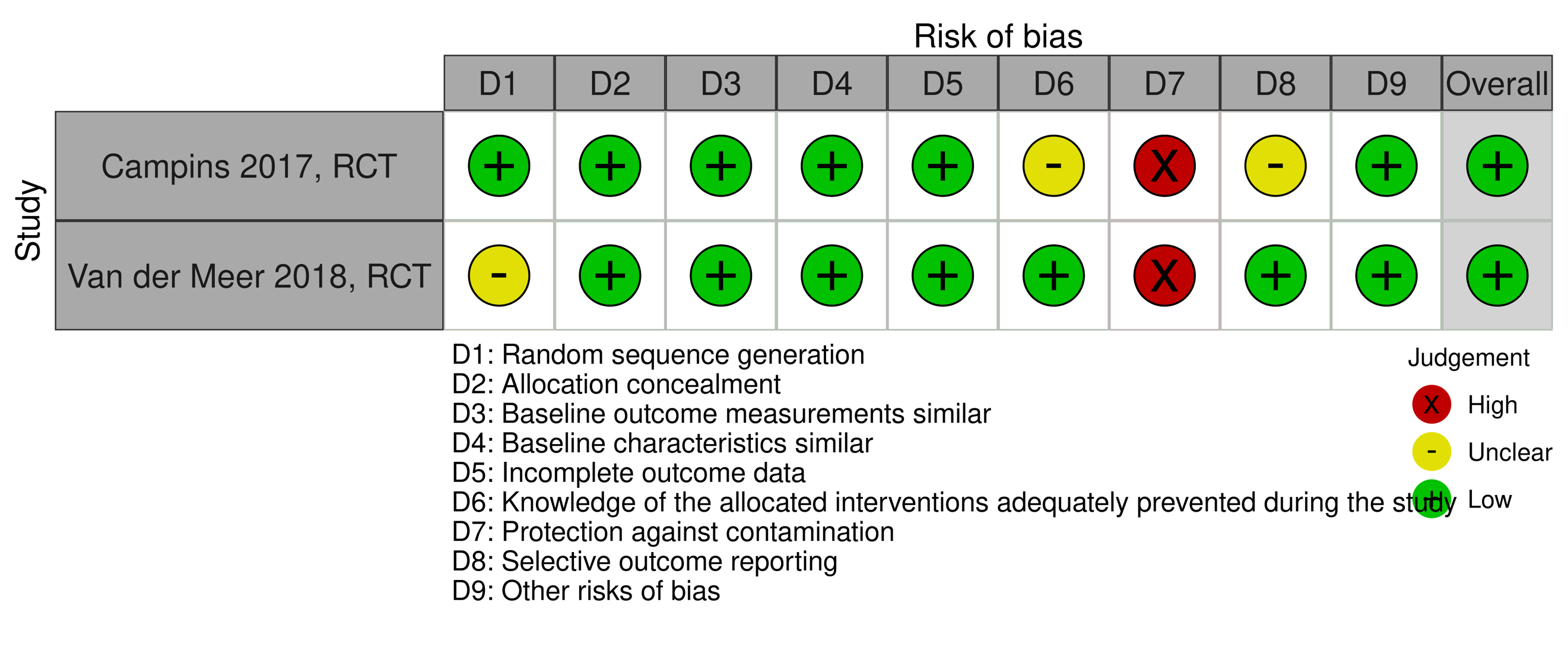

### 6. CHEC List for health-economic studies

|  | Campins (2019) | Cowper (1998) | Jodar-Sanchez (2015) | Malone (2000) | Noain (2017) |
| --- | --- | --- | --- | --- | --- |
| Is the study population clearly described? | Y | Y | Y | Y | N |
| Are competing alternatives clearly described? | Y | Y | Y | Y | N |
| Is a well-defined research question posed in answerable form? | Y | U | Y | U | U |
| Is the economic study design appropriate to the stated objective? | Y | U | Y | U | Y |
| Is the chosen time horizon appropriate to include relevant costs | Y | Y | Y | U | Y |
| Is the actual perspective chosen appropriate? | Y | Y | Y | Y | Y |
| Are all important and relevant costs for each alternative identified? | Y | Y | Y | Y | N |
| Are all costs measured appropriately in physical units? | U | U | U | Y | U |
| Are costs valued appropriately? | Y | U | U | Y | U |
| Are all important and relevant outcomes for each alternative identified? | N | N | Y | Y | N |
| Are all outcomes measured appropriately? | U | Y | Y | U | Y |
| Are outcomes valued appropriately? | U | N | Y | U | U |
| Is an incremental analysis of costs and outcomes of alternatives performed? | N | N | Y | N | N |
| Are all future costs and outcomes discounted appropriately? | N/A | U | N/A | N | N/A |
| Are all important variables, whose values are uncertain, appropriately subjected to sensitivity analysis? | U | N | N | Y | N |
| Do the conclusions follow from the data reported? | Y | U | Y | U | Y |
| Does the study discuss the generalizability of the results to other settings and patient/client groups? | U | N | N | U | N |
| Does the article indicate that there is no potential conflict of interest of study researcher(s) and funder(s)? | Y | U | Y | U | Y |
| Are ethical and distributional issues discussed appropriately? | N | N | U | N | N |

Y (yes), N (no), U (unclear), N/A (not applicable)
