## Additional File 4 for "The effectiveness and cost of integrating pharmacists within general practice to optimize prescribing and health outcomes in primary care patients with polypharmacy: A systematic review"

### Medication related problems

| **Author (Year)** | **Measure used, Baseline PIP Prevalence** | **Post-intervention PIP prevalence, defined study endpoint** | **Effect Sizes, significance** |
| --- | --- | --- | --- |
| **MAI** | | | |
| **Bryant (2011)**  **RCT** | Inappropriate medications as measured by mean MAI  Intervention, 5.1  Control, 4.5 | Mean MAI at 6 months  Intervention, 3.1  Control, 4.2 | Mean difference at 6 months*, 1.1 (95% CI -1.78 to -0.42, p=0.003) |
| **Hanlon (1996)**  **RCT** | Adjusted MAI score (per person)  Intervention, 17.7±0.6  Control, 17.6±0.6 | Adjusted MAI at 12 months  Intervention, 12.8±0.7  Control, 16.7±0.7 | 28% improvement (adjusted change score - 4.9) in the intervention group versus a 5% improvement (adjusted change score - 0.9) in the control group (P=0.0002) |
| **Taylor (2003)**  **RCT** | Number of inappropriate prescriptions as measured by MAI  Intervention, 210  Control, 207 | Number of inappropriate prescriptions at 12 months as measured by MAI  Intervention, 155  Control, 224 | Reduction in PIP intervention vs control  26% reduction in intervention vs 8% increase in control (calculated) |
| **STOPP/START Criteria** | | | |
| **Campins (2017)** | Drug changes based on DRPs as measured by STOPP/START criteria at 3 months (mean, SD)  *Intervention:*  Drug discontinuations, 1.27(1.29)  Dose adjustments, 0.96(1.15)  Drug substitutions, 0.49(0.8)  *Control:*  Drug discontinuations, 0.42(0.9)  Dose adjustments, 0.18(0.43)  Drug substitutions, 0.19(0.46) | Drug changes based on DRPs as measured by STOPP/START criteria at 12 months (mean, SD)  *Intervention:*  Drug discontinuations, 2.69(1.98)  Dose adjustments, 1.14(1.25)  Drug substitutions, 0.95(1.16)  *Control:*  Drug discontinuations, 2.05(1.91)  Dose adjustments, 0.37(0.65)  Drug substitutions, 0.64(0.85) | Drug discontinuations: 0.33 (p < 0.001)  Dose adjustments: 0.77 (p < 0.001)  Drug substitutions: 0.30 (p = 0.005) |
| **DRPs based on structural assessment by Cipolle** | | | |
| **Geurts (2016)**  **RCT** | DRPs based on structural assessment by Cipolle  Intervention, 394  Control, 47 | DRPs based on structural assessment by Cipolle at 12 months  Intervention, 208  Control, 4 | Reduction in PIP intervention vs control  47% vs 91% |
| **Kwint (2001)**  **RCT** | Number of DRPs (total) [mean per patient]  Intervention: 249 (4.5)  Control: 231 (4.4) | Number of DRPs (total) [mean per patient]  Intervention: 175 (3.2)  Control: 221 (4.2) | Reduction in PIP intervention vs control  29.7% vs 4.3% |
| **DBI** | | | |
| **Sloeserwij (2019)**  **nRCT** | DBI per patient, mean (SD)  Intervention, 0.48 (0.64)  Usual care, 0.53 (0.63)  Usual care plus, 0.78 (0.68) | DBI per patient, mean (SD) at 12 months  Intervention, 0.50 (0.63)  Usual care, 0.54 (0.64)  Usual care plus, 0.56 (0.67) | Adjusted treatment effect (95% CI) on lowering DBI  Intervention vs usual care, -0.02 (-0.07 – 0.02)  Intervention vs usual care plus, -0.01 (-0.06 – 0.04) |
| **VanDerMeer (2018)**  **RCT** | DBI [mean (SD)]  Intervention, 3.1 (1.0)  Control, 3.2 (1.0) | Proportion of patients with a decrease of DBI≥0.5 at 3 months  Intervention, 17.3%  Control, 15.9% | Not calculable |
| **Locally defined protocols** | | | |
| **Granas (1998)**  **RCT** | Locally defined DRPs  Intervention, 36.3%  Control, 34.1% | DRPs at 12 months  Intervention, 7.8%  Control, 11.6% | Adjusted ARR, 26% |
| **Krska (2001)**  **RCT** | Locally defined PCIs  Intervention, 1206 (100%)  Control, 1380 (100%) | PCIs at 3 months  Intervention, 256 (21.2%)  Control, 856 (60.7%) | RR = 0.35 95%CI (0.31-0.39) |
| **Vinks (2009)**  **RCT** | Locally defined DRPs in line with national guidelines [mean (SD)]  Intervention, 4.1 (2.4)  Control, 3.8 (2.6) | Mean number of DRPs at 4 months  Intervention, 3.29  Control, 3.62 | Mean difference -16.3% (95% CI -24.3, -8.3) |

Text highlighted in bold indicates a heading.

PIP (potentially inappropriate prescribing), MAI (medications appropriateness index), DRP (drug related problem), RCT (randomised controlled trial), nRCT (non-randomised controlled trial), DBI (drug burden index), SD (standard deviations), CI (confidence interval), ARR (absolute risk reduction), PCI (pharmaceutical care issue), RR (relative risk)
