## Additional File 5 for "The effectiveness and cost of integrating pharmacists within general practice to optimize prescribing and health outcomes in primary care patients with polypharmacy: A systematic review"

### Difference in number of medications

| **Author (Year)** | **No. of medicines at baseline (mean±SD, unless otherwise stated)** | **No. of Medicines post-intervention (mean±SD, unless otherwise stated)** |
| --- | --- | --- |
| **Bernsten (2001)** | Intervention 7.05 ± 2.51  Control: 6.97 ± 2.51 | Intervention: 7.14 ± 2.90,  Control: 7.00 ± 2.74 |
| **Britton (1991)** | Intervention 8.72 ± 3.54  Control: 8.52 ± 3.47 | Change post intervention:  Intervention: -0.21 ± 1.43  Control: 0.48 ± 1.18 |
| **Campins (2017)** | Intervention 10.79 ± 2.52  Control 10.91 ± 2.65 | Intervention n=10.03  Control n=10.91 |
| **Hanlon (1996)** | Intervention 7.6 ± 2.8 Control 8.2 ± 2.7 | Intervention 6.9 ± 2.6  Control 7.9 ± 3.3 |
| **Jodar-Sanchez (2015)** | Intervention, 7.76 (2.51)  Control, 7.32 (2.32) | Intervention, 7.48 (2.39)  Control, 7.25 (2.40)  Difference in reduction between groups  0.21 (0.06) 95% CI 0.092 – 0.335 |
| **Lenaghan (2007)** | Intervention 9.01 Control 9.85 | Intervention 8.68  Control 10.33  (mean difference in change in the number of items was -0.87 in favour of intervention) |
| **Taylor (2003)** | Intervention 6.3 ± 2.2 Control 5.7 ± 1.7 | Intervention 4.7 ± 2.0  Control 6.2 ± 2.0 |
| **Verdoorn (2019)** | No. Medications *(median, IQR):*  Intervention & control both n=9.0 (7.5-10.5) | Total no. of medications in intervention decreased with −0.32 medications after 6 months |
| **Vinks 2009** | Intervention 8.32 ± 2.5  Control 8.4 ± 2.3 | Intervention 8.32 ± 2.5  Control 8.4 ± 2.3 |

Text highlighted in bold indicates a heading.

SD (standard deviation), CI (confidence interval), IQR (interquartile range)
