## Additional File 6 for "The effectiveness and cost of integrating pharmacists within general practice to optimize prescribing and health outcomes in primary care patients with polypharmacy: A systematic review"

### 1. HRQoL: SF-36

| **Author (Year)** | **General Health** | | **Mental Health** | | **Physical Functioning** | | **Emotional role** | | **Physical role** | | **Vitality** | | **Bodily pain** | | **Social functioning** | |
| --- | --- | --- | --- | --- | --- | --- | --- | --- | --- | --- | --- | --- | --- | --- | --- | --- |
|  | **Baseline** | **Follow-up** | **Baseline** | **Follow-up** | **Baseline** | **Follow-up** | **Baseline** | **Follow-up** | **Baseline** | **Follow-up** | **Baseline** | **Follow-up** | **Baseline** | **Follow-up** | **Baseline** | **Follow-up** |
| **Bernsten (2001) [mean (SD)]** | I, 46.2 (20.4)  C, 46.5 (21.0) | Not reported as a total | I, 69.7 (21.5)  C, 70.1 (21.6) | Not reported as a total | I, 46.6 (26.1)  C, 47.4 (27.4) | Not reported as a total | I, 55.9 (44.6)  C, 65.2 (43.4) | Not reported as a total | I, 32.2 (4.4)  C, 32.9 (40.6) | Not reported as a total | I, 48.3 (22.9)  C, 49.6 (23.9) | Not reported as a total | I, 56.0 (27.8)  C, 55.4 (55.4) | Not reported as a total | I, 67.4 (27.8)  C, 68.1 (29.0) | Not reported as a total |
| **Bryant (2011)** | Not reported | | | | | | 13.4 unit difference in favour of control, p = 0.024 | | Not reported | | | | | | 7.7 unit difference in favour of control, p = 0.019 | |
| **Carter (2001)** | Not reported | | I, 70.78 (18.95)  C, Not reported | | Not reported | | | | I, 37.94 (41.68)  C, Not reported | | Not reported | | | | | |
| **Graffen (2004)** | Not reported | | P<0.0001 in favour of I | | Not reported | | | | | | P<0.009 in favour of I | | Not reported | | | |
| **Hanlon (1996) [mean (SD)]** | I, 34.9 (2.1)  C, 34.2 (2.1) | I, 37.4 (1.6)  C, 35.2 (1.7) | I, 61.0 (2.5)  C, 63.5 (2.5) | I, 61.1 (1.8)  C, 60.4 (1.8) | I, 48.0 (2.7)  C, 45.3 (2.7) | I, 44.0 (2.0)  C, 42.2 (2.0) | I, 73.0 (4.1)  C, 68.1 (4.1) | I, 66.4 (1.8)  C, 67.0 (3.9) | I, 38.3 (3.2)  C, 36.5 (3.2) | I, 38.6 (3.6)  C, 32.3 (3.7) | I, 31.7 (2.2)  C, 32.9 (2.2) | I, 36.5 (2.0)  C, 36.1 (2.0) | I, 45.0 (2.8)  C, 42.2 (2.8) | I, 43.6 (2.7)  C, 41.7 (2.7) | I, 56.4 (3.0)  C, 59.4 (3.0) | I, 57.4 (2.6)  C, 55.1 (2.7) |
| **Krska (2001)** | Not reported – stated no significant difference between domains | | | | | | | | | | | | | | | |
| **Sellors (2003)**  **(mean (95% CI))** | I, 62.2 (61.9–62.6)  C, 65.0 (64.8–65.2) | I, 60.5 (60.3–60.7)  C, 60.8 (60.6–61.0) | I, 75.2 (75.1–75.3)  C, 76.7 (75.8–77.6) | I, 74.2 (74.0–74.3)  C, 74.7 (74.7–74.8) | I, 55.6 (55.5–56.0)  C, 54.2 (48.0–54.4) | I, 55.0 (54.6–55.3)  C, 55.0 (54.8–55.2) | I, 71.8 (70.9–72.7)  C, 74.9 (74.5–75.2) | I, 66.4 (65.7–67.0)  C, 72.7 (72.1–73.2) | I, 53.8 (53.1–54.6)  C, 55.0 (54.5–55.5) | I, 48.5 (47.8–49.3)  C, 52.1 (41.6–42.6) | I, 53.8 (53.6–54.0)  C, 54.5 (53.0–56.0) | I, 52.7 (52.5–52.9)  C, 53.2 (53.1–53.3) | I, 60.5 (60.2–60.8)  C, 60.8 (60.6–61.0) | I, 56.6 (56.4–56.8)  C, 59.0 (58.8–59.2) | I, 79.2 (79.0–79.4)  C, 81.9 (81.8–82.0) | I, 75.4 (75.1–75.8)  C, 77.5 (77.3–77.7) |
| **Taylor (2003) [mean (SD)]** | I, 50.8 (19.5)  C, 49.9 (19.8) | I, 57.0 (19.6)  C, 50.1 (15.9) | I, 72.0 (17.4)  C, 69.0 (18.6) | I, 73.1 (21.2)  C, 72.3 (17.1) | I, 62.0 (29.4)  C, 61.9 (24.3) | I, 68.6 (24.0)  C, 56.1 (27.5) | I, 59.6 (44.7)  C, 69.4 (45.3) | I, 82.8 (36.4)  C, 65.8 (45.4) | I, 50.8 (42.2)  C,47.9 (42.8) | I, 68.2 (42.1)    C, 52.8 (42.2) | I, 47.0 (23.5)  C, 46.9 (24.1) | I, 55.6 (20.3)  C, 47.9 (20.2) | I, 60.0 (27.0)  C, 65.4 (23.0) | I, 68.5 (22.3)  C, 63.1 (25.8) | I, 70.6 (24.9)  C, 73.3 (26.6) | I, 77.8 (24.3)  C, 73.0 (28.2) |

Text highlighted in bold indicates a heading.

HRQoL (health related quality of life), SF-36 (short form 36), I (intervention), C (control), SD (standard deviation), CI (confidence interval)

### 2. Intervention effects on HRQoL - SF-36 Sorensen Trial

| **Sorensen (2004)** | **PCS** | | **MCS** | |
| --- | --- | --- | --- | --- |
|  | Baseline (mean (95% CI)) | Follow up (mean (95% CI)) | Baseline (mean (95% CI)) | Follow up (mean (95% CI)) |
|  | Intervention, 30.20 (27.87, 32.53)  Control, 32.35 (30.49, 34.21) | Intervention, 31.04 (29.04)  Control, 30.49 (29.08, 31.90) | Intervention, 51.22 (49.12, 53.32)  Control, 50.15 (48.62, 51.68) | Intervention, 48.67 (46.86, 50.49)  Control, 50.69 (49.06, 52.32) |

Text highlighted in bold indicates a heading.

HRQoL (health related quality of life), SF-36 (short form 36), PCS (physical component score), MCS (mental component score), CI (confidence interval)

### 3. Intervention Effect on HRQoL- EQ5D

|  | **Baseline** | **Follow-up** | **Summary** |
| --- | --- | --- | --- |
| **Campins (2017)** | Not reported | Not reported | Change in baseline score of -2.09 points in the intervention group and 0.67 points in the control group (p = 0.324) |
| **Leendertse (2013)** | Not reported | Not reported | *EQ5D*  No significant between group difference [0.16 (95% CI: 0.01–0.42)]  *EQ-VAS*  No significant between group difference [1.73 (95% CI:0.37–3.68)] |
| **Lenaghan (2007)** | *Mean EQ5D utility score*  Intervention, 0.62  Control, 0.57  *VAS*  Intervention, 63.7  Control, 65.2 | *Mean EQ5D scores at 6 months*  Intervention, 0.57  Control, 0.56  *VAS at six months*  Intervention, 63.8  Control, 68.3 | Utility score decreased in both groups, less in control group. No statistical difference. |
| **Van der Meer (2018)** | *EQ5D3L score*  Intervention, 48  Control, 61  *VAS [mean (SD)]*  Intervention, 6.6 (1.6)  Control, 6.8 (1.4) | *EQ5D3L score at follow up*  Intervention, 54  Control, 64  *VAS [mean (SD)]*  Intervention, 6.4 (1.6)  Control, 6.7 (1.5) | No difference |
| **Jodar-Sanchez (2015)** | *EQ5D3L mean utility score (SD)*  Intervention, 0.7148 (0.28)  Control, 0.6953 (0.31)  *VAS score (SD)*  Intervention, 65.44 (18.07)  Control, 63.22 (19.42) | *EQ5D3L mean utility score (SD)*  Intervention, 0.7677 (0.27)  Control, 0.6931 (0.32)  *VAS score (SD)*  Intervention, 70.46 (17.06)  Control, 62.29 (19.20) | Difference between intervention and control at follow-up,  Mean utility score (SD), 0.0550 (0.01) 95% CI (0.0306 – 0.0794),  VAS score, 5.87 (0.85) 95% CI (4.20 – 7.54) |
| **Varas-Doval (2020)** | Data not reported, change between intervention and control reported as univariate and multivariate analysis associated with uncontrolled health problems | Data not reported, change between intervention and control reported as univariate and multivariate analysis associated with uncontrolled health problems | *EQ5D Index Value Change per increase of 0.1 in EQ5D Index Value at any given time-point*  *Univariate*: -0.06 (-0.067 -0.052)  *Multivariate*: -0.04 (-0.05 - 0.03)  *EQ5D VAS Scale Change per increase of 10 in EQ5D VAS scale at any given time-point -Univariate*: 0.07 (-0.08 -0.06) *Multivariate*: -0.03 (-0.05 - 0.02) |
| **Verdoorn (2019)** | *EQ5D5L utility score [mean (SD)]*  Intervention, 0.73 (0.18)  Control, 0.74 (0.18)  *VAS [mean (SD)]*  Intervention, 68 (16)  Control, 70 (16) | *EQ5D5L utility score at follow up [mean (SD)]*  Intervention, 0.73 (0.20)  Control, 0.74 (0.18)  *VAS [mean (SD)]*  Intervention, 70 (16)  Control, 69 (15) | Improved over time in favour of intervention. +3.4 points in quality of life (95% CI 0.94 to 5.8; *p* = 0.006) |

Text highlighted in bold indicates a heading, text highlighted in italics indicates a subheading.

HRQoL (health related quality of life), EQ5D (European Quality of Life five dimensions), VAS (visual analogue scale), SD (standard deviation), CI (confidence interval)
