## Additional File 7 for "The effectiveness and cost of integrating pharmacists within general practice to optimize prescribing and health outcomes in primary care patients with polypharmacy: A systematic review"

### Change in reported ADEs

| **Study author (year)** | **Symptom Score** | **Baseline** | **Follow up, n (%)** | **Significance** |
| --- | --- | --- | --- | --- |
| **Jameson (2001)** | Improved (≥2 points)  Unchanged (+1 to – 1 points)  Worsened (≥2 points) | Symptom score  Intervention, 10.4  Control, 7.3 | *Intervention*  Improved 67 (54)  Unchanged 30 (24.2)  Worsened 27 (21.8)  *Control*  Improved 58 (40.2)  Unchanged 50 (34.7)  Worsened 46 (31.9) | The baseline adverse effects and symptoms score significantly different (p = 0.03).  Comparison of scores at the end of the study with baseline showed more patients improving and fewer worsening in the intervention group. Significant, p = 0.024 |
| **Leendertse (2013)** | Not reported | Not reported | *Number patients with one or more adverse events*  Intervention, 104 (28.6%) Control, 73, (23.5%) | OR 1.02 (95% CI: 0.94–1.08)] |
| **Sorensen (2004)** | Not reported | *ADEs, %*  Intervention, 36.9  Control, 34.9% | *ADEs, %*  Intervention, 9.3%  Control, 34.0% | Not calculable |
| **Taylor (2003)** | ADEs defined as medication misadventures, in which an iatrogenic incident occurs that may be attributable to “error, immunologic response, or idiosyncratic response and is always unexpected or undesirable to the patient. “ | Not reported | *Patients with at least one medication misadventure*  Intervention, 2.8%  Control, 3.0% | Not significant |
| **Van der Meer (2018)** | Sedative side effects and risk of falls. | *Sedative side effects, median IQR*  Intervention, 3.0 (5.0)  Control, 2.0 (4.0)  *Reported falls*  Not reported | *Sedative side effects, median IQR*  Intervention, 2.0 (3.0)  Control, 3.0 (4.0)  *Reported falls*  Intervention, 18 (30.5%)  Control, 15 (19.5%) | Not significant p=0.1 |

Text highlighted in bold indicates a heading, text highlighted in italics indicates a subheading.

OR (odds ratio), CI (confidence interval), ADE (adverse drug event), IQR (interquartile range)
