## Additional File 8 for "The effectiveness and cost of integrating pharmacists within general practice to optimize prescribing and health outcomes in primary care patients with polypharmacy: A systematic review"

### Health service utilization

|  | **Hospitalisations** | **ED attendance** | **Mortality** |
| --- | --- | --- | --- |
| **Bernsten (2001)** | Lower proportion in intervention reported hospitalisations than control (35.6% and 40.4%) but not statistically significant. | Not reported | Not reported |
| **Campins (2017)** | No significant between group change | No significant between group change | No significant between group change |
| **Graffen (2004)** | No significant between group change | Not reported | Not reported |
| **Krska (2001)** | No significant between group change | Decrease in emergency admissions:  Intervention = 74%  Control = 27%  No significant difference. | Not reported |
| **Leendertse (2013)** | Intervention n=6 (1.6%) medication related hospitalisation.  Control n=10 (3.2%) admissions.  No significant difference. (hazard ratio of 0.50, 95% CI: 0.12–1.59; p = 0.20). | Not reported | Not reported |
| **Lenaghan (2007)** | Intervention n=20 unplanned admissions  Control n=21 unplanned admissions  Non-significant reduction in admission of 8% (relative risk = 0*.*92, 95% CI 0.50 – 1.70, *P* = 0*.*80). | Not reported | Not reported |
| **Sloeserwij (2019)** | N=822 total of medication-related hospitalisations.  The adjusted rate ratio for medication‐related hospitalisations in the intervention group compared to usual care was 0.68 (95% CI 0.57–0.82) and compared to usual care plus 1.05 (95% CI 0.73–1.52) | Not reported | Not reported |
| **Taylor (2003)** | Year prior to study: No. hospitalisations  Intervention n=24, control n=11  During study year:  Intervention n=2, control n=11, (p=0.003) | Year prior to study: No. ED admittances  Intervention n=18, control n=6  During study year:  Intervention n=4, control n=6, (p=0.044) | Not reported |
| **Van der Meer (2018)** | No difference found between control arm and intervention arm in hospitalisation, with 9 (11.7%) vs 3 (5.1%) patients reporting unplanned hospital admission (p=0.149). | Not reported | Two patients died, one (1.2%) in control arm and one (1.3%) in intervention arm (p=0.732). |
| **Zillich (2014)** | Intervention did not signiﬁcantly reduce 60-day, all-cause hospitalization compared to the usual care group (Adjusted OR: 1.26, 95% CI: 0.89–1.77, p=0.19).  For patients in the quartile with lowest risk, intervention resulted in three times less hospitalizations at 60 days (Adjusted OR: 3.78, 95% CI: 1.35–10.57, p =0.01). | Not reported | Not reported |

Text highlighted in bold indicates a heading.

CI (confidence interval), ED (emergency department), OR (odds ratio)
