## Additional File 9 for "The effectiveness and cost of integrating pharmacists within general practice to optimize prescribing and health outcomes in primary care patients with polypharmacy: A systematic review"

### Clinical Physical Outcomes

|  | **Dyslipidaemia** | **Diabetes** | **Blood pressure** | **BMI** | **Renal function** | **Anticoagulation** |
| --- | --- | --- | --- | --- | --- | --- |
| **Carter (2001)** | Fasting lipid profile: significantly more in intervention group (p=0.021)  Total cholesterol (7.4 vs 17.7mg/dL, p=0.028) and LDL cholesterol (12.8 vs 23.4mg/dL, p=0.042) significantly lower in intervention group. | Significantly more HBA1C tests in intervention group (p=0.04).  No difference in HBA1C levels or reductions between control and intervention. | Not reported | Not reported | Not reported | Not reported |
| **Geurts (2016)** | Intervention patients had significantly increased HDL cholesterol after 1-year follow-up (79.8–76.8 mmHg; *p* = 0.008). | No significant difference in HbA1C levels. | Intervention patients had a significantly decreased diastolic blood pressure after 1-year follow-up (79.8–76.8 mmHg; *p* = 0.008). | No significant change in BMI. | No significant change in renal function. | Not reported |
| **Taylor (2003)** | The intervention group had a dramatic improvement in LDL cholesterol at 12 months, while the percentage of patients in the control group meeting LDL cholesterol goals declined. | Percentage of patients achieving the therapeutic goal increased from 23.1% to 100.0% in the intervention group during the 12-month period but decreased in the control group.  Significantly higher in intervention group than control. | At 12 months, intervention-group patients were significantly more likely than control patients to have targeted blood pressures.  Significant increase from baseline in the percentage of patients at goal in the intervention group. | Not reported | Not reported | At 12 months, all patients in the intervention group had INRs within the targeted range, but only 25% of control patients did. |

Headings are highlighted in bold.

LDL (low density lipoprotein), HBA1C (glycated haemoglobin), HDL (high density lipoprotein), BMI (body mass index), INR (international normalised ratio)
