## Additional File 10 for "The effectiveness and cost of integrating pharmacists within general practice to optimize prescribing and health outcomes in primary care patients with polypharmacy: A systematic review"

### Summary of results of included health-economic studies

| **Author (year)** | **Cost year and original currency** | **Cost categories** | **Incremental cost (intervention vs control)** | **Incremental effects (intervention vs control)** | **Cost-effectiveness** | **Uncertainty** |
| --- | --- | --- | --- | --- | --- | --- |
| **Campins (2019)** | 2012, Euro (€) | The analysis aimed to assess the reduction in pharmaceutical expenditure alongside the clinical trials due to the pharmacist intervention in primary care in relation to routine clinical practice. Monetary valuation of time: salary data available through the 2012 collective labour agreement.  Real cost in drug expenditure calculated | *Before-after differences reduction in drug expenditure (€), mean (SD), [95%CI]:*  Intervention, 233.75 (510.46), [169.83 – 297.67]  Control, 169.40 (527.35), [103.37 – 235.43]  P = 0.171, not significant  *Incremental drug expenditure of standard care over pharmacist intervention (€)*  64.30/patient per year | Not reported | Not reported | A sensitivity analysis was performed based on three theoretical  Scenarios;  Basal scenario (30 minutes pharmacist and 20 minutes physician time)  Optimistic scenario (20 minutes pharmacist and 15 minutes physician),  Conservative scenario (40 minutes pharmacist and 30 minutes physician per patient) |
| **Cowper (1998)** | 1991, US dollars ($) | *Fixed costs:*  Faculty time, pharmacist training, beeper  *Variable costs:*  Initial contact; chart review, patient interview, physician meeting, education  Follow-up; assessment, physician meeting, education. | Not reported | Not reported | *Cost-effectiveness ratio for intervention*  Mean change in MAI 4.0  *Ratio including drug costs:* (($1006 + $120) - $1096)/4.0, or $7.50/1 unit change in MAI.  *Ratio excluding drug costs:* ($120/4.0), or $30/1 unit change in the MAI | Not reported |
| **Jodar-Sanchez (2015)** | 2014, Euro (€) | Estimate the ICER of a medication review with follow-up for older adults with polypharmacy against standard care | Mean incremental total cost of –€250.51 ± 156.82 (95 % CI -558.17 to 57.14) | Mean incremental QALY of 0.0156 ± 0.004 (95 % CI 0.008–0.023) | If willingness to pay is between €30,000/QALY and €45,000/QALY, the probability of the MRF service being cost effective, compared with usual dispensing, is 100 %. | Performed a non-parametric bootstrapping with 5000 replications. Resulting 5000 ICER replicates plotted on cost-effectiveness plane and used to construct a cost-effectiveness acceptability curve. |
| **Noain (2017)** | 2014, Euro (€) | Requires estimates of the unit cost of supplying capacity and of the time required to perform the different activities necessary to provide a service.  Service provider cost, initial investment and maintenance costs. | Not reported | Not reported | Not reported- reported break-even analysis:  Break-even point varied from a minimum of 91 patients for scenarios 6 (highest estimated mark up and number of patients) to a maximum of 129 patients (scenario 1, lowest mark up and number of patients) | Not reported |
| **Malone (2000)** | 1998, US dollars ($) | Clinic visits  Drugs  Hospitalisations  Laboratory tests | Mean change in total costs in intervention group (before/after intervention): $1020  Mean change in total costs in control group (before/after intervention): $1313  Difference in change of mean -$293, p=0.06 | Not reported | Not reported | To determine the influence of assumptions made when assigning cost estimates to resources consumed, several sensitivity analyses were performed.  Primary analysis: costs assigned to medium intensity/30 min visits  Low intensity /15 minute  High intensity/60 minute |
| **Bernsten (2001)** | 1999, Euro (€) | Health-related resource use  Cost of intervention | Between-group analysis indicated that  there were no significant differences between the  total cost for control and intervention patients in  any country (Mann-Whitney, p > 0.05) | Not reported | Not reported | Not reported |
| **Britton (1991)** | 1991, US dollar ($) | Medication costs  Medication changes | *Intervention total cost savings:* €144.77 ($189)  *Control total cost savings:* costs increased by €651.61 (£850.67)  *Intervention total cost avoidance:* €798.67 ($1042.65) (based on figures of total cost increase in the control group: 315 x 3.31) | Not reported | Not reported | Not reported |
| **Jameson (2001)** | 2001, US dollar ($) | Medical costs  Drug costs | *Intervention:*  Drug baseline 1593 ± 1116  Post-enrolment 1657 ± 1068  Change 63 ± 771  *Control:*  Drug baseline 1582 ± 1016  Post-enrolment 1602 ± 1202  Change 20 ± 601  *Intervention:*  Medical baseline 3566 ± 6308  Post-enrolment 4105 ± 9838  Change 539 ± 9992  *Control:*  Medical baseline 3428 ± 8043  Post-enrolment 3741 ± 8026  Change 313 ± 10431 | Not reported | Not reported | Not reported |
| **Krska (2001)** | 2001, Sterling (£) | Medicine costs | *Intervention Medication Costs (mean, SD)*  Baseline: 39.23 (29.07)  Follow-up: 38.83 (29.60)  *Control Medication Costs*  Baseline: 42.80 (33.50)  Follow-up: 42.61 (31.84) | Not reported | Not reported | Not reported |
| **Sellors (2003)** | 2003, Canadian dollar (Can $) | Cost of medications  Cost of healthcare utilization | *Cost of medications*  Daily medication costs were also similar in the 2 groups: $5.01 versus $4.82 (p = 0.72) for total costs and $3.57 versus $3.76 (p = 0.78) for costs to the Ontario Drug Benefit Program.  *Cost of healthcare utilization*  Mean cost of health care resources per senior was $1281.27 in the intervention group and $1299.37 in the control group (p = 0.45) | Not reported | Not reported | Not reported |
| **Sloeserwij (2019)** | Not reported, Euro (€) | Healthcare costs  Medication costs | *Healthcare costs*  Primary care costs 1.08 (0.99–1.17) p=0.073  Secondary care costs 0.92 (0.65–1.29) p=0.622  Medication costs 1.04 (0.98–1.10) p=0.172  *Secondary healthcare costs related to hospitalisations,* no differences: adjusted ratio 0.82 (95% CI 0.64–1.06).  *Medication costs , median (IQR)*  Intervention  Pre: 841 (441–1581)  Post: 868 (450–1479)  Usual care  Pre: 857 (435–1532)  Post: 749 (400–1383) | Not reported | Not reported | Not reported |
| **Sorensen (2004)** | 2004; trial period, Australian dollar (Aus$) | Medication costs  Healthcare utilization costs | *Cumulative cost/patient over the 8 months from enrolment*  AUS$5730 (£2234) for the control group and AUS$5401 (£2105) for the intervention group.  Net cost saving per intervention patient (marginal cost benefit) was AUS$54 (~ £19) per patient relative to controls. | Not reported | *Incremental cost–effectiveness ratio in reducing ADEs and in improving DUSOI-A for the groups*  *ADEs:* €38.69 [AUS$69 (~ £24)]  *DUSOI-A:* €36.45 [AUS$65 (~ £23)] | Not reported |

Headings are highlighted in bold, subheadings highlighted with italics.

SD (standard deviation), CI (confidence interval), MAI (medicines appropriateness index), ICER (incremental cost effectiveness ratio), QALY (quality life years adjusted), MRF (medication review framework), IQR (inter quartile range), ADEs (adverse drug events), DUSOI-A (Duke Severity of Illness Scale)
